# A novel hyperactive *BCR::ABL1*^*e6a3*^ variant confers resistance to combined asciminib plus ponatinib therapy

**DOI:** 10.64898/2026.04.14.26349982

**Authors:** Valentina Nardi, Joshua Schwieterman, Sekhu Ansari, Zachary Kincaid, Mohammad Azhar, Tahira Yousuf, Noor Amir, Areeba Khan, Meenu Kesarwani, Scott Ryall, Andrew M. Brunner, Maria Reyes Capilla Guerra, Gabriel K. Griffin, Nicolas Nassar, George Q. Daley, Mohammad Azam

## Abstract

Despite considerable advances, the emergence of treatment resistance to tyrosine kinase inhibitors (TKIs) therapy remains a significant challenge in chronic myeloid leukemia (CML). Here, we report the first clinical case of resistance to combined ponatinib and asciminib therapy in a CML patient who relapsed with B lymphoblastic blast crisis. While at presentation the patient harbored the canonical *e13a2 BCR::ABL1* fusion, at relapse his disease harbored the T315I mutation together with a novel *e6a3 BCR::ABL1* fusion, arisen by internal deletion in the original translocated allele. Structural modeling and biochemical analyses demonstrated that deletion of exon 2–encoded residues of *ABL1* destabilizes the autoinhibited conformation, resulting in a hyperactive kinase with increased propensity for B-cell differentiation. Functional studies revealed that both *BCR::ABL1*^*e6a3*^ and *BCR::ABL1*^*e6a3/T315I*^ conferred resistance to ponatinib and asciminib, alone or in combination. *BCR::ABL1*^*e6a3*^ demonstrated enhanced sensitivity to active-state selective inhibitors dasatinib and bosutinib, whereas BCR::ABL1^e6a3/T315I^ remained resistant. Combined drug sensitivity assays showed that axitinib restored inhibitory activity when combined with ponatinib or asciminib. Strikingly, a combination of axitinib and asciminib with low dose ponatinib fully suppressed enzymatic activity of BCR::ABL1^e6a3/T315I^ and cellular proliferation. These data show that treatment with asciminib and ponatinib can select for mutations with notably elevated enzymatic activity, effectively targeted by an axitinib-based triple combination. These data highlight the remarkable mutability of the BCR::ABL1 kinase, including through novel isoforms and provides a strong rationale for the clinical assessment of a triple inhibitor combination as a strategy to overcome resistance to dual ponatinib and asciminib therapy.

## Introduction

Tyrosine kinase inhibitor (TKI) therapy has transformed the therapeutic paradigm of chronic myeloid leukemia (CML), enabling near-normal life expectancy in half of treated patients (1-5). Despite this, a substantial number of patients develop resistance, and outcomes remain poor in CML blast crisis Philadelphia chromosome-positive (Ph+) acute lymphoblastic leukemia (ALL) (6-8). In most cases, resistance arises from point mutations within the ABL1 kinase domain that either directly hinder drug binding or enhance kinase activity that render them refractory to inhibitors targeting the inactive kinase conformation (type-II inhibitor), such as imatinib, nilotinib, ribastinib, and ponatinib (9-14). Consequently, type-I inhibitors, including dasatinib and bosutinib, which target the active kinase conformation, have demonstrated efficacy in suppressing a broad spectrum of resistant mutations, although the gatekeeper variants remain refractory (10, 15-17).

Multiple ABL1 kinase inhibitors targeting ATP-site or allosteric site are in clinical use and when employed strategically, can effectively address resistance mediated by BCR::ABL1 mutations. Among these, ponatinib, an ATP-site inhibitor, and asciminib, an allosteric inhibitor targeting the myristoyl pocket, have shown significant activity against the T315I gatekeeper mutation (12, 18). While ponatinib has demonstrated meaningful clinical efficacy in patients with TKI-resistant disease, its broader application is constrained by dose-dependent toxicities. In addition, resistance frequently emerges through compound kinase domain mutations (i.e., two or more concurrent mutations (14), as well as through mutation-independent mechanisms(19). Importantly, compound mutants harboring the gatekeeper mutation, are refractory to all approved ABL1 inhibitors (12, 14, 20, 21). In contrast, asciminib monotherapy has demonstrated notable efficacy in both newly diagnosed (22-25) and TKI-resistant patients (26), with a favorable safety profile. Earlier, we demonstrated that destabilizing the active-state by dismantling the hydrophobic spine abrogates resistance conferred by the gatekeeper mutant (11). Both asciminib and ponatinib suppress the gatekeeper mutant at 10–30-fold higher concentrations than wild-type BCR::ABL1, likely by destabilizing the active conformation stabilized by the gatekeeper mutation. Subsequent studies confirmed that ponatinib and asciminib in combination synergistically inhibit both gatekeeper and ponatinib-resistant compound mutants (12, 27, 28). Building on these promising observations, combinatorial approaches in CML particularly in blast crisis patients are in clinical trials (NCT03595917, NCT03578367, NCT02081378). Considering the history of cancer treatment and the dynamic nature of the replicative genome to evolve under selective pressure, the emergence of resistance to combination therapies is inevitable.

Here, we present a clinical case in which resistance to combined ponatinib and asciminib treatment arose through the selection of a novel *BCR::ABL1*^*e6a3*^ fusion together with the gatekeeper mutation T315I. Expression of *BCR::ABL1*^*e6a3*^ and its variants in primary human cells revealed increased differentiation toward the B-cell lineage and conferred resistance to ponatinib and asciminib, both as single agents and in combination. A combination of axitinib and asciminib with low-dose ponatinib fully overcame BCR::ABL1^e6a3/T315I^ –mediated resistance in both murine and primary human cells. These findings provide compelling evidence that patients treated with ponatinib and asciminib may select for mutations driving hyperactive kinase activity, which can be suppressed by adding a gatekeeper-selective, conformation-tolerant inhibitor such as axitinib in combination regimens.

## Methods

### *BCR::ABL1* quantitative RT-PCR for *e13a2* and *e14a2* transcripts

The clinically validated QuantideX qPCR BCR-ABL IS (Asuragen, Texas) is an FDA cleared assay to quantify *BCR::ABL1 e13a2* and *e14a2* transcript on the international scale with deep clinical sensitivity of 0.002% IS (MR4.7).

### Targeted RNA sequencing assay

The Oncomine Myeloid Assay GX v2 Assay (Thermo Fisher scientific, Massachusetts) is a clinically validated ultrarapid [https://doi.org/10.1182/blood-2024-208059] next generation sequencing tumor profiling test investigating specific single nucleotide variants, insertions/deletions and gene fusions in genomic DNA and RNA using an Ion Torrent Integrated Sequencer, as per manufacturer instructions. The Archer FusionPlex assay with custom primers (IDT, Iowa) is a clinically validated targeted anchored multiplex based RNA sequencing assay to detect gene fusions in hematological and solid tumor malignancies (29).

### Targeted DNA sequencing assay

The Rapid Heme panel assay (New England Biolabs, Massachusetts) is a clinically validated custom targeted next generation sequencing of DNA test investigating single nucleotide variants, insertions/deletions and copy number changes in 88 genes, with a validated LOD of 3% VAF.

### Nanopore targeted sequencing of *BCR* and *ABL1*

To assess the presence of the breakpoint in the genomic DNA, two independent long range PCR reactions were performed using two different sets of custom forward and reverse primers spanning the exon 5 of the *BCR* gene to exon 4 of *ABL1*. PCR reactions were conducted as described in the PCR Protocol for LongAmp Hot Start Taq DNA Polymerase (NEB, Cat. No. M0534) following manufacturer’s instructions (1st set of primers, forward primer sequence: ATGCCTTGATGCCGTTCAGA; reverse primer sequence: CTCGTACACCTCCCCGTACT; 2nd set of primers, forward primer sequence: ACCATTTCAGCGAATGGGGT; reverse primer sequence: CTTCAAGGTCTTCACGGCCA). PCR products were purified using QIAquick PCR Purification Kit for PCR Cleanup (Qiagen, Cat. No. 28106) and quality control of the PCR product was conducted by checking DNA amount using high sensitivity dsDNA qubit kit (Cat. No. Q33231) and DNA size using TapeStation (Cat. No. 5067-5366). Then, both products followed library preparation according to the Oxford Nanopore Technologies (ONT) ligation sequencing kit (Cat. No. SQK-LSK114). Once prepared, DNA libraries were loaded onto two independent flongle flow cells and sequenced for 24 hours with active base calling (Cat. No. FLO-FLG114). All sample preparation and sequencing methods were conducted according to ONT manufacturer’s instructions. Bam files were inspected to look for genomic breakpoints involving both genes, which were found to be in chr22: g.23614371 for *BCR* gene and chr9: g.133729793 for *ABL1* gene. Coordinates were confirmed in both PCR products.

### Chromosomal microarray

Chromosomal microarray analysis is a clinically validated assay performed on DNA extracted from bone marrow using the Applied BiosystemsTM CytoScanTM HD Accel assay, a high density, whole genome microarray with 2,772,571 probes. The array consists of 2,029,441 non-polymorphic regions and 743,130 single-nucleotide polymorphisms (SNPs), with an average genome-wide spacing of 1.1 kb. Chromosome Analysis SuiteTM software was used to compare, in silico, the hybridization pattern of a patient specimen against a pooled reference sample set.

### IGH clonality testing

This clinically validated DNA assay consists of a polymerase chain reaction technique using primers conjugated with fluorescent dyes that hybridize to conserved framework (regions 1,2 and 3) and joining regions of the immunoglobulin heavy chain gene (InVivoScribe Technologies, BIOMED, San Diego, CA). The PCR products are then analyzed by capillary gel electrophoresis.

Plasmid construction, mutagenesis, cell proliferation assays, structural predictions, and immunoblotting were carried out as previously described (9, 11, 30). A detailed methodology is provided in the Supplemental methods section.

### Statistical Analysis

Statistical analyses were conducted using Prism software v9.0 (GraphPad Software, La Jolla, CA, USA). The median survival was calculated by log-rank test. For in vitro studies, statistical significance was determined by the two-tailed unpaired Student’s t-test. A p value < 0.05 was considered statistically significant.

## Results

### Patient clinical history

A patient in his 70’s with chronic myeloid leukemia (CML) characterized by the *BCR::ABL1 (e13a2)* fusion transcript was initially treated with imatinib (**Figure 1A**). After eight years, the patient achieved a deep molecular response (DMR; ≥4–4.5 log reduction), which was sustained for two additional years before treatment was discontinued. The patient remained in treatment-free remission (TFR) for 2.5 years, but routine testing detected molecular relapse. The patient was initially treated with dasatinib, but due to gastrointestinal toxicity, therapy was switched to imatinib, to which he initially responded. However, six months later, he progressed to a B-lymphoid blast crisis (B-ALL) with high *BCR::ABL1*^*e13a2*^ >50%IS and the emergence of an imatinib-resistant P-loop mutation, G250E (**Figure 1A**). Imatinib was discontinued, and hydroxyurea was started, but failed to control disease progression. The patient was then treated with ponatinib plus prednisone, which produced only modest activity without remission. He was subsequently switched to combined ponatinib and asciminib therapy that resulted in a rapid hematologic response with deep molecular remission (DMR) within three months (**Figure 1B**). However, at six months of treatment patient relapsed with B-ALL. Targeted RNA sequencing with two different assays identified a novel in-frame *BCR::ABL1* transcript involving *BCR* exon 6 and *ABL1* exon 3, *e6a3* (**Supplemental Fig. 1A**) while *e13a2* was detected at 0.0123% IS (Figure 1B). IGH Clonality assay showed the same clonal rearrangement between the initial *e13a2* blast crisis and the e6a3 one (**Supplemental Fig. 1B**). Targeted DNA sequencing identified some similar mutations and some evidence of clonal evolution (**Supplemental table 1**). Chromosomal microarray analysis and Nanopore long-read sequencing confirmed an internal deletion in the *BCR::ABL1* allele (**Supplemental Fig. 1 C and D**), resulting in the formation of a novel *e6a3* fusion transcript harboring the gatekeeper mutation T315I (**Figure 1C, Supplemental Fig. 1)**. Unfortunately, the patient succumbed to disease progression two weeks after relapse.

**Figure 1.**
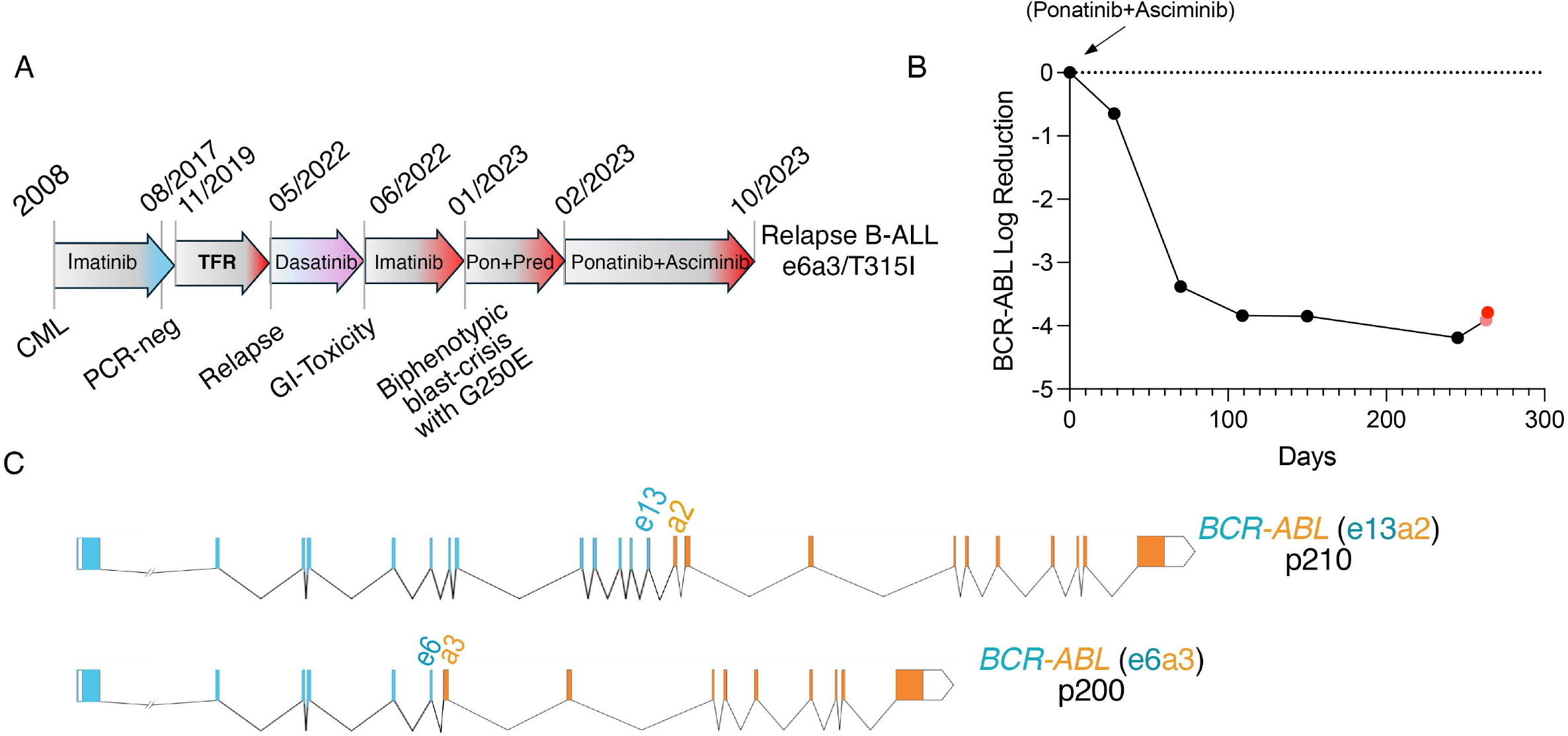
Clinical course of a CML patient that developed resistance to combined ponatinib and asciminib therapy. **A**. Schematic timeline showing treatment response to imatinib, achievement of treatment free remission, and subsequently developed resistance to combination of ponatinib and asciminib treatment. **B**. Quantitative PCR analysis showing depletion of *BCR::ABL1* transcript levels (%IS) during treatment. Patient achieved remission within three months with four log reduction in *BCR::ABL1* transcripts; loss of remission indicated by increased *BCR::ABL1* levels (red dots). **C**. Shown are the primary structures of *BCR::ABL1*^*e13a2*^ (exon 13 of *BCR* fused to exon2 of *ABL1* gene) at the time of diagnosis and of *BCR::ABL1*^*e6a3*^ (exon 6 of *BCR* fused to exon 3 of *ABL1* gene).

### *BCR::ABL1*^*e6a3*^ encodes a hyperactive kinase

Several BCR::ABL1 protein isoforms result from distinct translocation breakpoints within the *BCR* gene, while all retain *ABL1* exons 2–11. Translocations involving *BCR* exons 13 and 14 produce the BCR::ABL1 p210 fusion protein while rearrangements involving exon 1 generate the p190 isoform. The p210 isoform is predominantly associated with CML, whereas p190 is more common in B-ALL. Differences in kinase activity, substrate specificity, and the type of progenitor cell in which the translocation occurs are thought to underlie the distinct pathological outcomes of these isoforms (31-36). The novel *e6a3* translocation is predicted to encode a ∼200 kDa BCR::ABL1 fusion protein with a molecular weight intermediate between the canonical p190 and p210 isoforms. Structural modeling showed that deletion of ABL1 exon 2– encoded residues removes a substantial portion of the SH3 domain, which is essential for stabilizing the SH2 domain at the kinase C-lobe and for properly positioning the SH2-kinase linker to allow SH3 docking at the N-lobe (37) (**Figure 2A–C**). Consistent with this interpretation, mutations within CAP, SH3, and SH2 domains are recurrently selected under therapeutic pressure from imatinib and other type-II inhibitors (9, 10, 38). Absence of exon 2 of ABL1 was also shown to confer resistance also to asciminib (37). In previous work, we identified multiple imatinib‐resistant variants arising within the CAP (E38 to E60) (39) and SH3 domain (N64 to D71) (39) encoded by exon 2 **(Figure 2C**). To confirm whether these variants disrupt kinase autoinhibition and enhance enzymatic activity, we introduced these mutations into wild-type ABL1. As reported earlier, G2A variant of ABL1b (ABL1^G2A^) displayed increased autophosphorylation of Y393 (11, 40), (**Figure 2D**). As envisioned, imatinib‐resistant variants from exon 2 exhibited increased autophosphorylation, confirming that these substitutions destabilize the autoinhibited state and promote kinase activation (**Figure 2D)**. Modeling predictions using AlphaFold2 suggested that deletion of exon 2–encoded residues promotes stabilization of the active kinase conformation (**Figure 2E,F**). In this model, I145 from the SH2 domain inserts into a hydrophobic pocket in the kinase N-lobe (comprised of residues M244, Y257, V268, A269, V270 and Y312) where residue Y215 packs above the I145 to further stabilize this interaction (**Figure 2E and F**). Further structural analysis revealed that this hydrophobic pocket is positioned above the gatekeeper threonine, such that residue I313 links the SH2-binding pocket with both the gatekeeper and the regulatory hydrophobic-spine assembled in the active state (11)(**Figure 2G and H**). Mutations within this hydrophobic network are frequently detected and confer TKI resistance (9, 10). Notably, substitutions within the regulatory spine or at I313 that introduce smaller amino acids (e.g., alanine) disrupt this interaction and cause enzymatic inactivation (9, 11). In contrast, enhanced hydrophobic interaction by variants, such as T315I/M or M244V/I stabilize the kinase in its active-state conformation (9, 11). Both regulatory and catalytic spines are coupled through a hydrophobic connectivity in the C-lobe that collectively modulates kinase conformational dynamics (41-43). These analyses predicted that BCR::ABL1^e6a3^ would be stabilized in an active conformation. Expression of BCR::ABL1^e6a3^ produced a ∼200 kDa protein with markedly enhanced auto- and transphosphorylation activity compared to the p190 and p210 variants (**Fig. 2I–J)**. Notably, BCR::ABL1 e6a3 harboring gatekeeper mutations showed ∼1.7-fold (T315I) and ∼2.5-fold (T315M) increases in total tyrosine phosphorylation, along with 13- and 15-fold higher autophosphorylation, respectively, relative to BCR::ABL1 p210 (**Supplemental Fig. 2**). In contrast, BCR::ABL1 p190 exhibited a six-fold increase in autophosphorylation but ∼50% lower transphosphorylation activity compared to p210 (**Fig. 2I–J, Supplemental Fig. 2**). Together, these results indicate that the e6a3 translocation encodes a hyperactive BCR::ABL1 kinase, and that co-occurring gatekeeper mutations further amplify its enzymatic activity.

**Figure 2.**
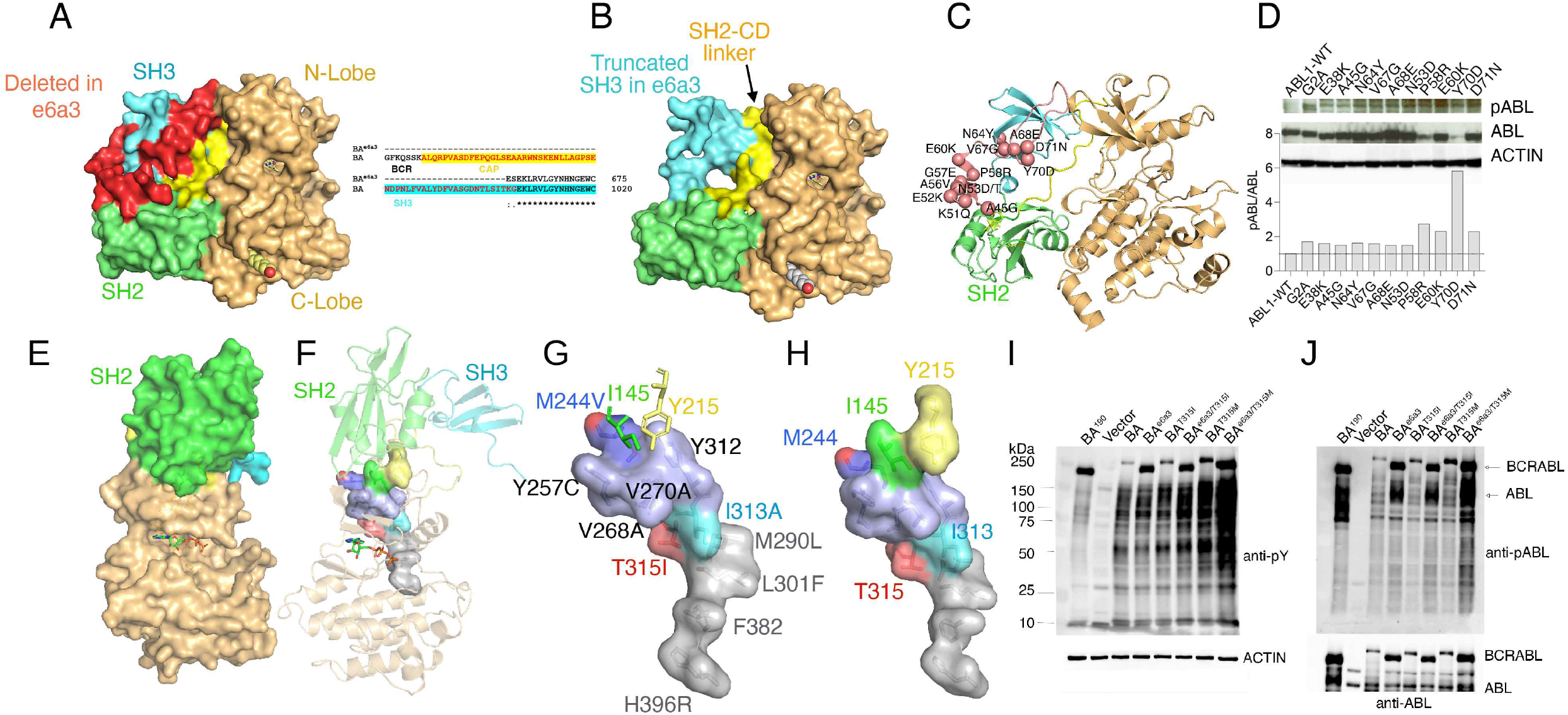
*BCR::ABL1*^*e6a3*^ encodes a hyperactive kinase. **A**. Surface representation of the autoinhibited ABL1 kinase highlighting key structural elements encoded by exon 2 (red) that are essential for maintaining the autoinhibited conformation. **B**. Surface representation of the autoinhibited ABL1 structure illustrating the truncated SH3 domain (cyan) that lacks exon 2-encoded residues. **C**. Ribbon depiction of the autoinhibited ABL1 structure showing exon 2–encoded residues (red spheres) identified in our imatinib-resistance mutagenesis screen (Azam et.al, 2003) **D**. Immunoblot demonstrating activation of ABL1 kinase activity (increased autophosphorylation at Y393) in the G2A mutant (lacking myristoylation, impaired autoinhibition) and exon 2–derived imatinib-resistant variants. **E**. Surface depiction of the active ABL1^e6a3^ isoform predicted by AlphaFold2, showing SH2 docking onto the N-lobe of the kinase domain. **F**. Ribbon view of the predicted active ABL1^e6a3^ conformation emphasizing SH2 engagement with its N-lobe binding pocket. **G**. Surface view of the SH2-binding pocket on the N-lobe formed by M244, Y257, V268, V270, and Y312. I145 (green sticks) inserts into this pocket, while Y215 (yellow sticks) stabilizes the interaction. Notably, the SH2-binding pocket connects to the regulatory spine (gray surface) through T315 (gatekeeper) and I313. **H**. Surface depiction illustrating how I145 is tightly packed between Y215, M244, and Y257, forming a hydrophobic anchor that stabilizes SH2 docking and active state conformation. **E**. Immunoblots showing total phosphotyrosine levels from the whole cell extracts of HEK293T cells expressing BCR::ABL1 and its variants. Total actin levels shown as the loading control (bottom panel). **F**. Immunoblots showing phospho-ABL1 (phospho-Y412) levels from the whole cell extracts of HEK293T cells expressing BCR::ABL1 and its variants. Total ABL1 levels shown below. Note increased total phosphotyrosine levels and enhanced autophosphorylation with BCR::ABL1^e6a3^ with an additive increase in ABL activation by the gatekeeper mutations, T315I and T315M. Remarkably, the T315M variant shows a stronger activation compared to T315I. Quantified values for each blot are provided in supplemental figure 2.

### BCR::ABL1^e6a3/T315I^ is resistant to combined ponatinib and asciminib

Ponatinib and asciminib both preferentially bind to inactive ABL1 kinase conformations, albeit at distinct sites that underlies their synergistic inhibitory activity (12, 18). While dual inhibition that stabilizes the inactive kinase conformation can suppress most resistant variants including compound mutations (28, 44), mutations favoring the active-state conformation may still emerge under this therapy. Consistent with this, BaF3 cells expressing the hyperactive BCR::ABL1^e6a3^ kinase exhibited ∼10-fold and ∼5000-fold resistance to ponatinib and asciminib, respectively (**Figure 3A and B)**. While the combination of both drugs largely suppressed this resistance, the IC_50_ remained modestly elevated compared with native BCR::ABL1 (**Figure 3C, Supplemental Table 2**). Despite the noted synergy of combined ponatinib and asciminib on BCR::ABL1 (12) and BCR::ABL1^T315I^ (28), the BCR::ABL1^e6a3/T315I^ variant demonstrated marked resistance, exhibiting a 13-fold reduction in sensitivity to asciminib and a 20-fold reduction to ponatinib, even under combination treatment. (**Figure 3C**). Similarly, the frequently observed ponatinib-resistant gatekeeper mutant T315M, whether alone or combined with e6a3, showed near-complete resistance to ponatinib and asciminib combination (**Figure 3C, Supplemental Table 2)**. Consistently, the hyperactive BCR::ABL1^e6a3^ kinase showed resistance to ABL1 type-II inhibitors imatinib, nilotinib, and ribastinib/DCC2036 (**Supplemental Figure 3A-E and Supplemental tables 3-4**) while exhibiting hypersensitivity to type-I inhibitors, dasatinib and bosutinib (**Figure 3D-I; and Supplemental Tables 5–8**). In contrast, gatekeeper mutations in both native BCR::ABL1 and the e6a3 variant conferred complete resistance to both dasatinib and bosutinib, most likely by sterically hindering inhibitor binding (**Figure 3D and G**). Notably, the combination of either type I or type II inhibitors with asciminib failed to overcome resistance mediated by BCR::ABL1^e6a3/T315I^ (Figure 3 and Supplemental Figure 3). These results suggested that gatekeeper-selective ABL1 inhibitors targeting the active kinase conformation, such as VX-680 and XL228, may effectively suppress BCR::ABL1^e6a3/T315I^. Expectedly, both compounds demonstrated inhibitory activity against T315I and e6a3/T315I variants, but their limited therapeutic windows precluded further clinical studies (45) (**Supplemental Fig. 4 and Supplemental Tables 9 and 10**). These findings confirm that the BCR::ABL1^e6a3/T315I^ variant observed at relapse was selected under combined asciminib and ponatinib therapy. Studies with inhibitor combinations suggest that combining a gatekeeper-selective, conformation-tolerant ABL1 inhibitor may overcome BCR::ABL1^e6a3/T315I^ –mediated resistance.

**Figure 3.**
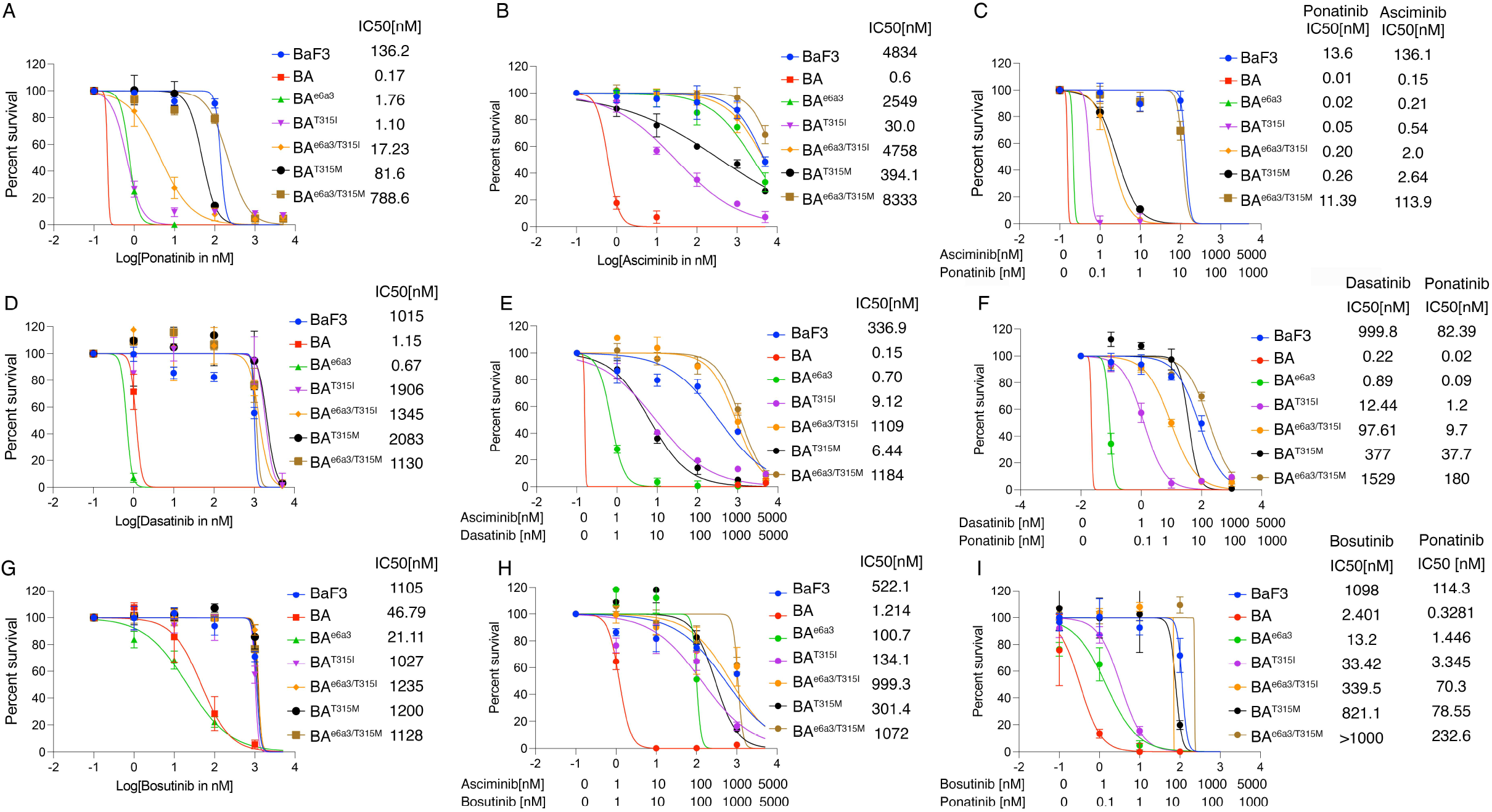
BCR::ABL1^e6a3/T315I^ confers resistance to ponatinib and asciminib combination treatment. Sigmoidal curve showing the viability of BaF3 cells expressing BCR::ABL1^e13a2^ (BA) and BCR::ABL1^e6a3^ variants treated with increasing concentrations of ABL inhibitors. IC50 values for each cell line is indicated in parentheses. **A**. Dose-dependent inhibition curve showing increased IC_50_ for ponatinib against BCR::ABL1^e6a3^ (∼10-fold higher than native BCR::ABL1^e13a2^, labeled as BA). The IC_50_ for ponatinib against BCR::ABL1^e6a3/T315M^ is approximately 100-fold higher than for native BA. Similarly, BCR::ABL1^T315M^ and BCR::ABL1^e6a3/T315M^ exhibit ∼80-fold and ∼780-fold increases in IC_50_, respectively, relative to native BA. **B**. Dose-dependent inhibition curve showing increased IC_50_ values for BCR::ABL1^e6a3^, BCR::ABL1^e6a3/T315I^, BCR::ABL1^T315M,^ and BCR::ABL1^e6a3/T315M^. **C**. Dose-dependent inhibition curve for the combination of asciminib and ponatinib showing increased IC_50_ values for BCR::ABL1^e6a3^ (∼2-fold), BCR::ABL1^e6a3/T315I^ (>20-fold), BCR::ABL1^T315M^ (>20-fold), and BCR::ABL1^e6a3/T315M^ (>1000-fold) relative to native BA. **D-I**. Dose-dependent inhibition curves showing reduced IC_50_ values for the type I ABL inhibitors dasatinib (**D**) and bosutinib (**G**) against BCR::ABL1^e6a3^, indicating that BCR::ABL1^e6a3^ stabilizes the active kinase state. In contrast, the gatekeeper mutations, whether in the BA or BCR::ABL1^e6a3^ configuration, confer resistance to both dasatinib and bosutinib, alone or in combination with asciminib (**E** and **H**) or ponatinib (**F** and **I**). Data shown are from three independent experiments.

**Figure 4.**
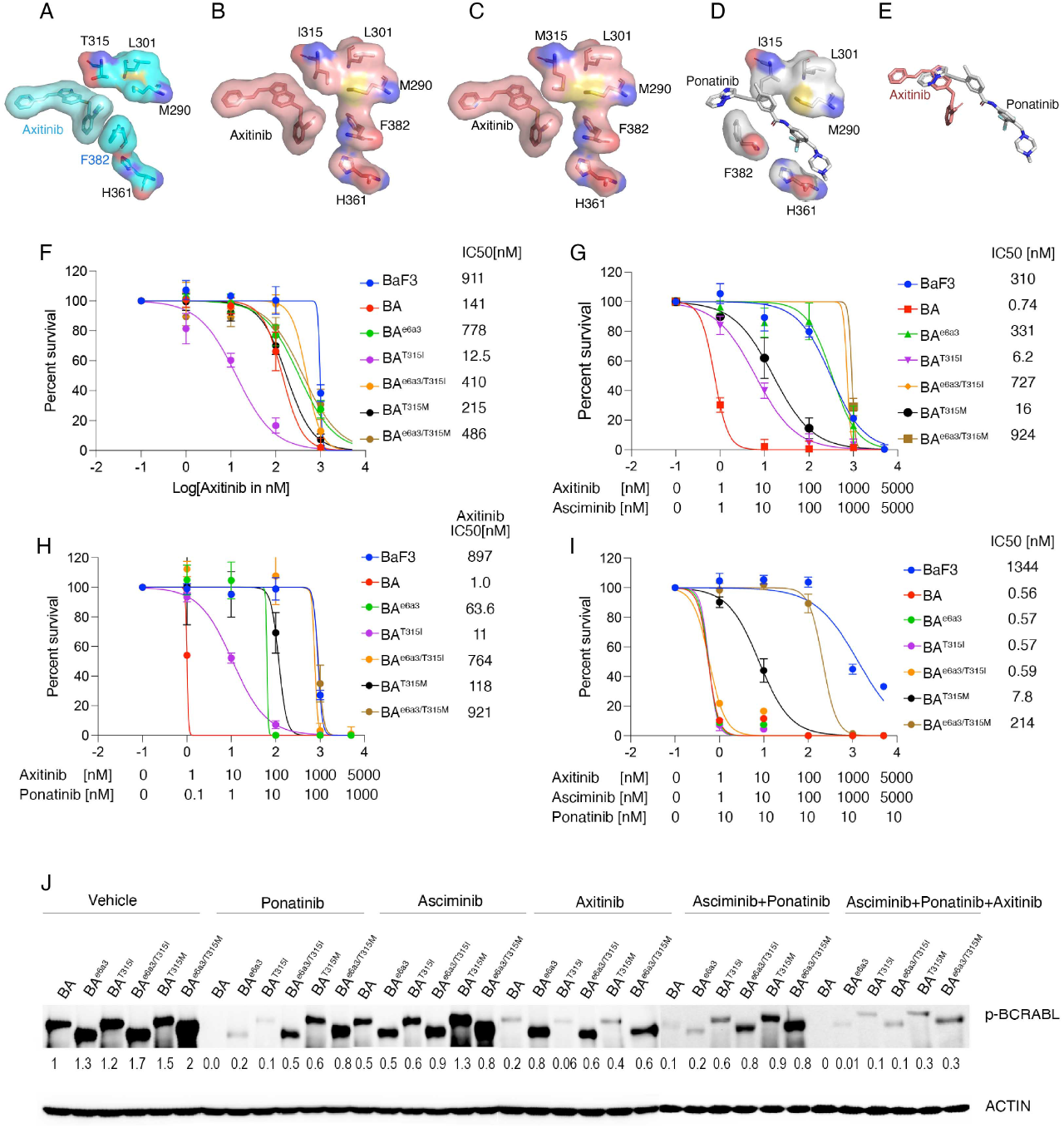
A combination of axitinib with ponatinib and asciminib overcomes resistance conferred by BCR::ABL1^e6a3/T315I^. **A**. Crystal structure of axitinib bound to wild-type ABL1 kinase in the DFG-out (inactive) conformation, where the regulatory spine is disrupted. **B**. Structure of axitinib bound to ABL1-T315I in the DFG-in (active) conformation with a stabilized regulatory spine. **C**. Model of axitinib binding to the ABL1 active site with the T315M mutation. The methionine substitution may introduce a modest steric clash with axitinib, whereas substitution with isoleucine does not affect binding. **D**. Ponatinib bound to the active site of ABL1 kinase in the DFG-out (inactive) conformation. **E**. Structural overlay showing that ponatinib and axitinib bind distinct sites within the active site with minimal overlap in their binding modes. **F**. Sigmoidal curve showing selective inhibition of BCR::ABL1^T315I^ by axitinib, with only modest inhibitory activity against native BCR::ABL1. In contrast, the BCR::ABL1^e6a3^ variant, alone or in combination with gatekeeper mutations, confers resistance to axitinib. **G**. Sigmoidal curve showing that axitinib combined with asciminib potently suppresses native BCR::ABL1 and its gatekeeper-resistant variants (T315I and T315M), whereas the BCR::ABL1^e6a3^ variant remains resistant. **H**. Sigmoidal curve demonstrating that axitinib combined with ponatinib suppresses BCR::ABL1^e6a3^ and potently inhibits native BCR::ABL1 and its gatekeeper mutants, while gatekeeper mutations in the BCR::ABL1^e6a3^ background remain resistant. **I**. Sigmoidal curve showing that the combination of axitinib and asciminib with low-dose ponatinib (10 nM) exhibits similar inhibitory activity against BCR::ABL1 and BCR::ABL1^e6a3^ variants harboring T315I mutations. In contrast, T315M variants display higher IC_50_ values, which can be suppressed at higher drug concentrations, albeit with a notable therapeutic index. **J**. Immunoblots showing phospho–BCR::ABL1 levels in whole cell extracts treated with ponatinib, asciminib, and axitinib alone or in combination. Notably, the triple combination of ponatinib, asciminib, and axitinib potently inhibits BCR::ABL1 and BCR::ABL1^e6a3^ variants. Total actin is shown as a loading control in the bottom panel. Representative data is from three independent experiments. The data were normalized to the vehicle control of the respective cell line.

### Axitinib in combination with ponatinib and asciminib suppresses BCR::ABL1^e6a3/T315I^

We identified axitinib as a conformation-tolerant inhibitor for further evaluation, given its ability to bind wild-type ABL1 in the inactive DFG-out conformation while engaging the T315I mutant in the active DFG-in conformation (**Figure 4A–B; Supplemental Figure 5A–F**) (46). Structural modeling further suggested that axitinib may retain activity against the T315M variant, albeit with reduced potency due to steric hindrance imposed by the methionine substitution (**Figure 4C**). Notably, ponatinib and axitinib occupy distinct yet partially overlapping regions within the ATP-binding pocket (**Figure 4D–E**), raising the possibility that dual ATP-competitive inhibition could result either in competitive displacement or, alternatively cerate space for cooperative binding. Consistent with prior reports, axitinib exhibited ∼10-fold greater sensitivity against BCR::ABL1^T315I^, whereas the e6a3 variants conferred relative resistance (**Figure 4F**).

**Figure 5.**
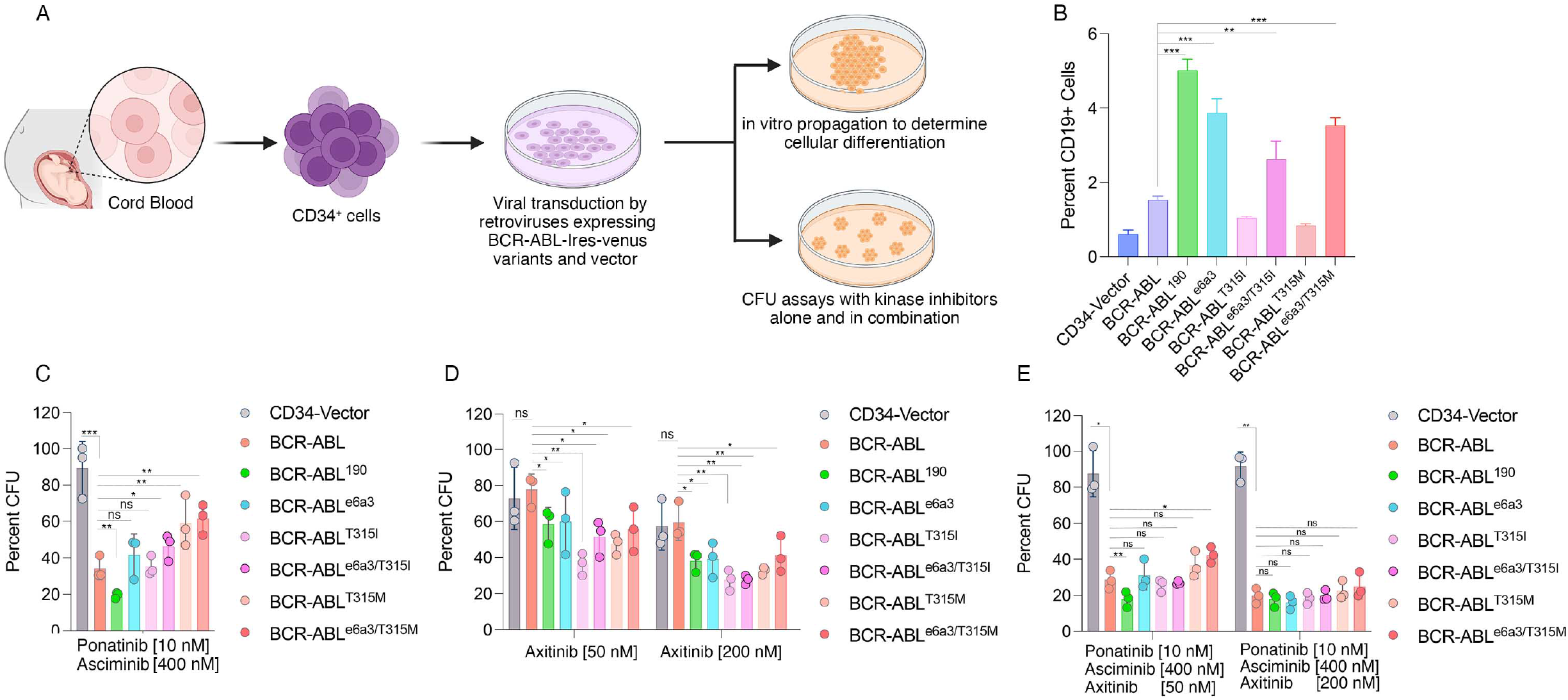
A combination of axitinib with ponatinib and asciminib overcomes resistance conferred by BCR::ABL1^e6a3^ and its gatekeeper variants in human cells. **A**. Schematic diagram showing isolation of CD34^+^ cells from human cord blood, followed by in vitro propagation for lineage differentiation and colony-forming unit (CFU) assays. **B**. Bar graph showing the percentage of CD19 positive cells derived from CD34 cells expressing BCR::ABL1 variants. Similar to BCR::ABL1^p190^, which is commonly associated with B-ALL, expression of BCR::ABL1^e6a3^ and its gatekeeper variants causes enhanced CD19 expression. **C**. Bar graph showing the percent CFUs from the cells treated with ponatinib and asciminib in combination. Notably, BCR::ABL1^e6a3^ and its variants, unlike BCR::ABL1^p190^ which is more sensitive to inhibition relative to native BCR::ABL1, confer resistance to dual ponatinib and asciminib treatment. **D**. Bar graph showing CFU percentages following treatment with axitinib at 50 nM (left panel) and 200 nM (right panel). Axitinib at 50 nM potently inhibits CFUs derived from BCR::ABL1^T315I^ cells, with modest activity against the other BCR::ABL1 variants and no activity against the native BCR::ABL1. At 200 nM, axitinib significantly suppresses CFUs for all BCR::ABL1 variants except the native BCR::ABL1. **E**. Bar graph showing that axitinib (50 nM) combined with ponatinib and asciminib significantly reduces CFUs for most variants except BCR::ABL1^T315M^ and BCR::ABL1^e6a3/T315M^. Strikingly, the triple combination with axitinib at 200 nM achieves potent inhibition of all BCR::ABL1 variants while preserving >90% CFU formation from the normal CD34^+^ cells, exhibiting a favorable therapeutic index. Representative data are from two independent experiments, presented as mean ± S.D. (n = 3); * = p < 0.05, ** = p < 0.01, and n.s. = not significant.

Expectedly, T315M variant, exhibited a higher IC_50_ relative to T315I, likely due to reduced binding space introduced by the methionine side chain (**Figure 4 C and F)**. These data confirm that axitinib is a T315I-selective inhibitor and can bind the inactive kinase conformation, as demonstrated by its activity against wild-type BCR::ABL1. We next evaluated axitinib in combination with other ABL1 inhibitors to identify agents capable of synergizing with it to inhibit BCR::ABL1^e6a3^ and its variants. The combination of axitinib with ponatinib produced strong synergistic inhibition of BCR::ABL1^e6a3^ and its gatekeeper variants, with combination index (CI) values ranging from 0.1 to 0.28 while its combination with other ATP-site ABL1 inhibitors lacked consistent inhibitory activity (**Figure 4G–H, Supplemental Figure 6, and Supplemental Tables 11–16**). In contrast, the combination of axitinib and asciminib demonstrated synergistic activity only against BCR::ABL1^e6a3^, whereas e6a3 variants harboring gatekeeper mutations remained insensitive to the combination (**Figure 4G**). These observations suggest that combining the allosteric inhibitor asciminib with ponatinib and axitinib may effectively sensitize the BCR::ABL1^e6a3/T315I^ variant. Remarkably, a combination of axitinib with asciminib and low-dose ponatinib (15 nM) completely inhibited BCR::ABL1 ^e6a3/T315I^, achieving sub-nanomolar IC_50_ values (∼0.5 nM) (**Figure 4I, Supplemental Table 17**). Notably, this triple combination also suppressed BCR::ABL1 T315M and BCR::ABL1 e6a3/T315I, albeit with higher IC_50_ values, while maintaining a significant therapeutic window relative to parental BaF3 cells (**Figure 4I**). Next, we examined total tyrosine phosphorylation and BCR::ABL1 autophosphorylation, which confirmed that the cytotoxic activity of the triple drug combination is mediated by direct on-target inhibition of BCR::ABL1 kinase activity (**Figure 4J, Supplemental Fig. 7**).

### Combined inhibition with axitinib, ponatinib, and asciminib overcomes resistance mediated by BCR::ABL1^e6a3^ and its gatekeeper variants in human CD34^+^ cells

Because enhanced kinase activity of BCR::ABL1 ^p190^ has been implicated in B-ALL transformation (31), we hypothesized that BCR::ABL1 e6a3 and its variants, like p190, would similarly promote B-cell expansion. Expression of BCR::ABL1^e6a3^ and its gatekeeper variants in human cord blood–derived CD34^+^ cells, similar to BCR::ABL1 p190, led to increased expansion of CD19^+^ B-cells compared with BCR::ABL1 p210 (**Figure 5A–B**). Under extended culture conditions, e6a3 and its variants consistently maintained elevated CD19 expression (30–40%) (**Supplemental Figure 8**), supporting the notion that the emergence of e6a3 at relapse in the patient likely drove B-ALL transformation. Next, to assess therapeutic relevance, we performed CFU assays with and without TKIs. As expected, BCR::ABL1^e6a3^ and its variants exhibited resistance to the ponatinib–asciminib combination while axitinib alone was effective only against BCR::ABL1^T315I^ (**Figure 5C-D**). Remarkably, the triple combination including 50 nM axitinib substantially reduced CFU numbers in cells expressing e6a3 and e6a3/T315I, while the e6a3/T315M variant showed modest resistance (**Figure 5E**). Interestingly, increasing axitinib to 200 nM, which is physiologically achievable based on PK data, suppressed all variants while preserving the clonogenic potential of parental CD34^+^ cells, demonstrating a favorable therapeutic window (**Figure 5E**). These results demonstrate that an axitinib-based triple combination overcomes resistance mediated by BCR::ABL1^e6a3^ and its gatekeeper variants while sparing normal hematopoietic progenitors.

## Discussion

The emergence of resistance to TKI therapy remains a critical clinical challenge. Acquisition of point mutations in the kinase domain that either directly impair drug binding or promote kinase conformations with reduced inhibitor affinity are the primary mechanisms of resistance. Inhibitors targeting inactive conformations, type-II (Imatinib, nilotinib and ponatinib) and allosteric type-IV (asciminib) select a broad spectrum of mutations compared to type-I inhibitors (dasatinib and bosutinib)(9, 10, 12-14, 47). Mutations that activate the kinase are often selected under type-II and IV inhibitors, as they not only reduce drug binding but also promote cancer cell survival and proliferation. Treatment with potent inhibitors or multiple TKIs can select for compound mutations which could pose significant challenge.

Protein kinase activity is governed by two conserved intramolecular hydrophobic networks that dynamically assemble in response to ATP binding or substrate engagement, forming a “*hydrophobic switch*” (41, 43). Our analysis has identified a novel hydrophobic connectivity stretching from the SH2 docking site within the N-lobe to the myristate binding pocket of the C-lobe (**Supplemental Figure 9 A-C)**. The first hydrophobic network we characterized, known as the regulatory spine (R-spine), is a hallmark of active kinases and plays a crucial role in substrate recognition, binding and drug resistance (11). Subsequently, a second hydrophobic network, the *catalytic spine* (C-spine), engaged in ATP binding has been defined, and mutations from this region have been reported to confer drug resistance **(Supplemental Figure 9 D-F**)(9, 42, 43). Assembly of the R-spine defines activation, whereas assembly of the C-spine poises the kinase for catalysis. Our modeling analysis suggests a third hydrophobic network within the C lobe of the kinase, which we dub the *hydrophobic girdle* (**Supplemental Figure 9 A-C)**. Importantly, allosteric interactions of the SH2 domain with the kinase domain have been shown to switch the activation loop from a closed to a fully open conformation, enabling activation loop autophosphorylation and supporting active kinase conformations (48).

A significant number of residues comprising the hydrophobic girdle have been reported in imatinib-resistant patients and identified in our prior screens (V338G, V339A/G, V379A, M351T/I, F359C, I360F, F486S, and M472I). However, the underlying mechanism has remained unclear. This new girdle model suggests that these mutations likely disrupt girdle assembly or alter its dynamics, thereby affecting imatinib binding. In the autoinhibited state of ABL1, SH2 docking at the C-lobe likely facilitates myristate binding and disrupts the hydrophobic girdle by displacing F497. In the active state, SH2 docking at the N-lobe promotes assembly of hydrophobic girdle to support the R- and C-spines rendering the kinase for catalysis (**Supplemental Figure 9E–H**). Thus, the SH2 domain plays a dual regulatory role: reinforcing autoinhibition at the C-lobe and promoting activation at the N-lobe. This framework explains how myristate and asciminib inhibit ABL1 allosterically and how activating mutations, including M244 and the gatekeeper residue, transmit allosteric signals by stabilizing R- and C-spines through the hydrophobic network. Overall, this model offers a mechanistic framework for SH2-mediated regulation of kinase conformational dynamics and activation states by modulating hydrophobic switches, especially the hydrophobic girdle at the C-lobe and the regulatory spine through M244 and the gatekeeper residue, and clarifies how displacement of the SH2 domain by e6a3 drives kinase activation and resistance.

The two major BCR::ABL1 isoforms, p210 and p190, drive distinct disease phenotypes, with p210 primarily characterizing CML and p190 classically linked to B-ALL or lymphoid blast crisis. Patients in advanced lymphoid or myeloid blast crisis exhibit poor responses to TKI therapy, a pattern reminiscent of the TKI-resistant phenotype observed in leukemic stem cells (LSCs) where growth factor signaling is known to enhance the apoptotic threshold (30). We suspect that, like p190, the e6a3 mutation promotes B-ALL that restores protective growth factor signaling, thereby raising the apoptotic threshold and providing an additional layer of resistance. Given that the combination of ponatinib and asciminib completely inhibited both BCR::ABL1^T315I^ and BCR::ABL1^e6a3/T315I^ mutants at higher drug concentrations, it indicated that these variants remain susceptible to enforced stabilization in the autoinhibited state. Similarly, BCR::ABL1^T315M^ was fully inhibited at elevated drug concentrations while its e6a3/T315M variant remained completely resistant, indicating that the exceptionally high enzymatic activity of this mutant drives resistance. Nonetheless, enhanced selectivity of type-I inhibitors, dasatinib and bosutinib, toward BCR::ABL1^e6a3^ while their failure against the gatekeeper variants due to steric hinderance highlight the need to develop gatekeeper-selective type-I inhibitors to overcome resistance driven by hyperactive kinases.

Higher doses of ponatinib and asciminib may partially overcome BCR::ABL1^e6a3/T315I^–mediated resistance, but off-target toxicities limit the clinical utility of this approach. Our data suggest that adding axitinib to this combination could help overcome resistance from both compound mutants, BCR::ABL1^e6a3/T315I^ and BCR::ABL1^e6a3/T315M^. Mechanistically, axitinib is a conformation-tolerant inhibitor that binds wild-type ABL1 in its inactive state (DFG-out), while targeting the T315I variant in its active state (DFG-in). Its greater potency against T315I (>10-fold relative to wild-type) is derived from increased complementarity with the hydrophobic surface introduced by the isoleucine substitution (46). In contrast, substitution with methionine causes a modest reduction in space within the ATP-binding pocket, which appears to restrict drug access and lead to resistance. Importantly, axitinib occupies a distinct region within the ATP-binding site compared to ponatinib, supporting the potential for cooperative binding interactions. It is tempting to speculate that axitinib or ponatinib binding induces conformational changes that create additional space for combinatorial orthosteric or allosteric binding. Alternatively, given that the BCR::ABL1 holoenzyme exists in a tetrameric complex, ponatinib and axitinib could engage distinct ATP-binding sites in adjacent kinases within the complex, together cooperatively stabilizing the inactive conformation within oligomers that constitute the BCR::ABL1 holoenzyme (49). Confirmation of any of the above speculations would require high-resolution structural studies of BCR::ABL1 holoenzyme to elucidate the mechanistic basis of noted synergies between orthosteric inhibitors. Notably, the triple combination of axitinib and asciminib with lower concentrations of ponatinib effectively suppressed BCR::ABL1^e6a3/T315I^, whereas inhibition of BCR::ABL1^e6a3/T315M^ required higher axitinib doses, which may be achievable through dose escalation. Importantly, the triple combination improved the therapeutic index in both murine Ba/F3 cells and primary human CD34^+^ cells, indicating that the adverse effects associated with ponatinib could be mitigated while retaining efficacy against resistant variants of BCR::ABL1. We posit that kinome modulation by axitinib and asciminib, combined with low-dose ponatinib, preferentially suppresses BCR::ABL1-driven signaling while sparing normal cells. Future proteomic studies will elucidate the molecular basis underlying the selectivity of this triple combination.

In summary, we report a novel mechanism of tyrosine kinase inhibitor (TKI) resistance involving a deletion event that results in loss of critical functional domains in ABL1 that encodes a hyperactive kinase, BCR::ABL1 ^e6a3^, resistant to ponatinib and asciminib combination therapy. Importantly, the prevalence of this phenomenon remains undetermined, potentially due to its under-recognition by standard molecular diagnostic approaches that primarily target common *BCR::ABL1* isoforms (*e13a2, e14a2* and *e1a2*). Given that axitinib is already approved for renal cell carcinoma with minimal adverse events, our study provides a strong rationale for clinical evaluation of axitinib in combination with ponatinib and asciminib to overcome broad-spectrum BCR::ABL1 drug-resistant mutations.

## Data Availability

All data produced in the present work are contained in the manuscript

## Acknowledgements

This study was supported by grants to MA from the National Cancer Institutes at NIH (RO1CA211594) and (RO1CA250516).

## Author Contribution

M..A, G.Q.D., and M..K wrote the manuscript. M.A., Z.K., M.K., Z.K., J.M, J.S., S.A., A.R., A.A, R.S., A.K., N.S., and N.A. performed the experiments and analyzed the data. M. A performed the in-silico docking studies. V.N, G.G.,M.D.L.R. and S.R. performed experiments on patient samples. V.N. and N..N edited the manuscript. All authors read and approved the manuscript.

