## Supplemental methods, figures (1-9) and tables (1-17) for "A novel hyperactive *BCR::ABL1*^*e6a3*^ variant confers resistance to combined asciminib plus ponatinib therapy"

### Supplemental materials

#### Supplemental Methods:

##### Plasmids and inhibitors

Retroviral plasmids expressing oncogenic kinases (BCR::ABL1, BCR::ABL1 T315I) were described earlier (1-3). Plasmids BCR::ABL1e6a3 and BCR::ABL1e6a3/T315I were built with the Q5 mutagenesis kit from New England Biolabs (NEB) according to manufacturer's instruction using the primers (e6a3FP: 5'- GTGAAAAGCTCCGGGTCTTAGG-3' and e6a3RP: 5'-TTTCCAGAGAGTTCTTGGTCGTTGG-3'. Kinase inhibitors including imatinib, nilotinib, dasatinib, ponatinib, asciminib, ribastinib, VX680, XL228, and axitinib were purchased from AdooQ Biosciences.

##### Cell Lines

BaF3, an IL3-dependent hematopoietic pro-B cell line, was transduced for stable expression of BCR::ABL1 mutants. Retroviral production and transduction were performed as described24,26. Cells were grown in RPMI supplemented with 10% FCS except supplemented with or without 10 ng/ml mouse IL3 as described (4, 5).

##### Cell-viability and western-blotting assays

Sensitivity to kinase inhibitors was determined by dose-escalation cell proliferation assays using BaF3 expressing *BCR::ABL1* or its variants. Ten thousand IL3-independent BaF3 cells stably expressing *BCR::ABL1* or its variants were plated in triplicate in 96-well plates in 0.1 ml RPMI medium containing 10% FCS with varying inhibitor concentrations. After 60 hours, cell viability was assessed using WST-1 reagent (Takara Bio Inc, USA). Absorbance, A450 nm, was averaged and plotted against inhibitor concentration as a best-fit sigmoidal curve, using the Prizm (GraphPad). The concentration resulting in 50% maximal inhibition was reported as the cellular IC50. Combination index (CIIC50) was calculated using the formula as described earlier (6, 7)combination index (CI)= (D1)c /(D1)a + (D2)c/(D2)a. (D1)a and (D2)a represent the concentrations of each drug aloneat IC50, while (D1)c and (D2)c were the concentrations of drugs in combination at IC50. CI < 1,= 1 and > 1 indicated synergism, additivity, and antagonism, respectively.

Four to six million cells were suspended in lysis buffer followed by three short bursts of sonication. Composition of lysis buffer have been described earlier. Lysates were separated by 10% SDS-PAGE and transferred to supported nitrocellulose membrane (BioRad) and probed with the following antibodies Anti- Phospho Tyrosine (santa cruz biotechnology) , anti Phospho ABL1 and anti- total ABL1 kinase( cell signaling technology. All primary antibodies were used at dilutions as recommended by the manufacturer. Anti-mouse or anti-rabbit IgG HRP conjugate secondary antibodies (GE Healthcare) were used at a 1:5000 dilution. HRP conjugated antibodies β-Actin (13E5) (#5125) from Cell Signaling Technology, Inc was used. Immunoblots were developed using SuperSignal West Dura Extended Duration Substrate (Thermo Scientific) followed with scanning on ChemiDocTM touch Imaging system (Bio-Rad). All western blots were replicated twice.

##### Structure prediction using AlphaFold2

Structural prediction of ABL1 e6a3 was performed using AlphaFold2 (v2.3) with default neural network weights and databases, following the standard pipeline as described earlier. The highest-ranked model was selected based on global predicted local distance difference test

(pLDDT) and predicted aligned error (PAE). Multiple-sequence alignments were generated automatically from UniRef90, MGnify, and environmental sequence databases using the default AlphaFold pipeline. No manual curation of MSA depth was performed. The top-ranked model was used for downstream evaluation. Disordered or low-confidence regions (pLDDT < 50) were retained but interpreted cautiously. Molecular graphics and visualization was made with PyMOL.
Isolation of human CD34+ cells, ex vivo culture and CFU assays.
Human CD34+ cells from normal cord blood were isolated using magnetic beads as described(ref). Hundred thousand CD34+ cells from both normal and leukemic samples were seeded with and without FLT3 inhibitors in SFEM media reconstituted with 100 ng/ml (SCF, FLT3LG, TPO, and GM-CSF) and 10 ng of IL3 and IL6. At day six, cells were harvested and stained for Annexin V and PI along with CD34 and CD38 cell surface markers as described earlier. Labeled cells were analyzed by FACS and results were analyzed using flowjo. Five thousand CD34 cells were seeded with and without kinase inhibitors in methocult (stemcell technology) in triplicate. Colonies were enumerated at day fourteenth and presented as percent relative to vehicle treatment (considered as 100%).

#### Supplemental Figure legends:

**Supplemental Figure 1. Detection of BCR::ABL1e6a3 rearrangement presumably through an intragenic deletion event.**
A. Schematic representation of the BCR exon6::ABL1 exon 2 fusion transcripts detected by Anchored Multiplex PCR (Archer Heme fusionplex, IDT, Iowa) and corresponding translation sequence.
B. IGH VDJ clonality testing with Framework I (top two electropherograms) and II (bottom two electropherograms) primers showing identical clonal rearrangements between the e13a2 lymphoid blast crisis and the e6a3 lymphoid blast crisis.
C. Results of chromosomal microarrays. Microarray analysis identified a 101.9kb deletion of 9q and a 23.0kb deletion of 22q which partially encompassed ABL1 and BCR, respectively. Based on the breakpoints identified, this finding results in a deletion extending from intron 1 to intron 2 (just prior to exon 3) of ABL1 and from intron 6-15 of BCR.
D. Intronic breakpoints in BCR downstream of exon 6 and ABL1 upstream of exon 3 detected by targeted DNA sequencing using Oxford Nanopore technology.

**Supplemental Figure 2. Gatekeeper mutations and BCR::ABL1$^{e6a3}$ translocation encode hyperactive kinase.**
**Bar graphs showing quantification of western blot from Figure 2 I**
**A. Total phosphotyrosine levels normalized to actin from the total cells extracts of HEK293T cells expressing BCR::ABL1 and its variants.**
**B. Showing phospho-BCR::ABL1 levels normalized to ABL1.**

**Supplemental Figure 3. BCR::ABL1$^{e6a3/T315I}$ confers resistance to type-II ABL inhibitors.**
**Dose dependent** inhibition curve showing the viability of BaF3 cells expressing BCR::ABL1 variants treated with increasing concentrations of imatinib (A), nilotinib (B), nilotinib and asciminib (C), ribastinib (D), and ribastinib with asciminib (E).

**Supplemental Figure 4. BCR::ABL1$^{e6a3/T315I}$ confers resistance to type-I gatekeeper selective ABL inhibitors.**

**Dose dependent** inhibition curve showing the viability of BaF3 cells expressing BCR::ABL1 variants treated with increasing concentrations of XL228 (A), Xl228 with asciminib (B), VX680 (C) and VX680 with asciminib (E).

**Supplemental Figure 5. Axitinib binds wild type ABL in inactive state while it targets T315I mutant in inactive conformation.**
A. Ribbon depiction of ABL1-wildtype kinase domain bound with axitinib (PDB: 4WA9). Axitinib and residues comprising the regulatory spine are shown as cyan sticks while activation loop is shown in golden color.
B. Structure $ABL1^{T315I}$ kinase domain bound with axitinib (PDB: 4TWP). Axitinib and residues comprising the regulatory spine are shown as pink sticks, activation loop shown in golden color.
C. Overlay of axitinib bound active and inactive conformations. Note, only the structures of DFG motifs are different. In inactive conformation, phenylalanine of DFG motif adopts DFG-out conformation where aspartate of DFG motif moves away from the active site and phenylalanine occupies the position of aspartate that results in broken spine activation loop is extended and open conformation like it was seen with active conformation (called as SRC-like inactive conformation) that seemingly targeted by axitinib in the wild type of ABL. In contrast, T315I mutant stabilizes regulatory-spine by adopting the DFG-in conformation that favors effective axitinib binding.
D-F. Close-up view of the active sites of ABL kinase showing the interaction of axitinib with ABL1-wildtype (D), $ABL1^{T315I}$ (E), and overlay of both structures (F). Note, the changes in the position of Phe382 in active (pink) and inactive state (cyan).

**Supplemental Figure 6. Axitinib in combination with ABL TKI exhibit variable response.**
**Dose dependent** inhibition curve showing the viability of BaF3 cells expressing BCR::ABL1 variants treated with increasing concentrations of axitinib in combination with nilotinib (A), ribastinib (B), XL228 (C), dasatinib (D), bosutinib (E) and VX680 (F).

**Supplemental Figure 7. A combination of axitinib with ponatinib and asciminib overcomes resistance conferred by $BCR::ABL1^{e6a3/T315I}$.**
Immunoblots showing total phospho tyrosine levels from the whole cell extracts of cells expressing BCR::ABL1 variants treated with ponatinib, asciminib, and axitinib alone or in combination. Notably, the triple combination of ponatinib, asciminib, and axitinib potently inhibits BCR::ABL1 mediated total phosphotyrosine levels. Total actin is shown as a loading control in the bottom panel. Representative data is from three independent experiments.

**Supplemental Figure 8. Expression of $BCR::ABL1^{e6a3}$ induces CD19 expression.**
Graph showing the percentage increase in CD19-positive cells during a three-week in vitro culture period. Notably, $BCR::ABL1^{p190}$ and $BCR::ABL1^{e6a3}$ variants induce a significant increase in the proportion of CD19-positive cells. Representative data is from two independent experiments presented as mean ± S.D. (n = 3); *p < 0.05, **p < 0.01, ***$p < 0.001$**, and n.s. = not significant.**

**Supplemental Figure 9. Hydrophobic modules involved in modulating the conformation and enzyme activity.**
A. Ribbon representation of $ABL1^{e6a3}$ in the active conformation. The SH2 domain is docked against the kinase N-lobe, stabilizing the regulatory-spine (R-spine; red), catalytic-spine (C-spine; green), and the hydrophobic girdle within the C-lobe (cyan).

B. Surface depiction of the hydrophobic girdle supporting the R- and C-spines. Notably, I145 (green) from the SH2 domain inserts into the N-lobe SH2-docking pocket formed by M244, A269, V270, Y257, and Y312. This SH2-docking pocket connects to the R-spine through the gatekeeper residue T315 and, whereas A270 links the pocket to the C-spine via V268 and F317.
C. Orthogonal view (90° rotation) highlighting the spatial organization of the hydrophobic girdle together with the R- and C-spines in active state.
D. Ribbon representation of the autoinhibited ABL1 structure, in which the SH2 domain docks against the C-lobe.
E. Inactive kinase conformation illustrating disruption of the regulatory spine (R-spine; red) and hydrophobic girdle (cyan). The myristoyl-group is shown in yellow.
F. Orthogonal (90°) view of the inactive kinase highlighting the spatial arrangement of the disrupted R- and C-spines and the hydrophobic girdle.
G. Cartoon representation of the autoinhibited ABL1 structure with dismantled regulatory-spine and hydrophobic girdle.
H. Cartoon representation of the active kinase structure, where the SH2 domain is lodged on top of the N-lobe, facilitating assembly of the hydrophobic girdle and regulatory spine.
I. A hypothetical model showing the BCR::ABL1 holoenzyme depicted as tetramer.

Supplemental Table 1. Comparison of Genomic Findings in the Two Blast Crises

Supplemental Tables 2-17. Table showing IC50 values and combination index of ABL inhibitors.

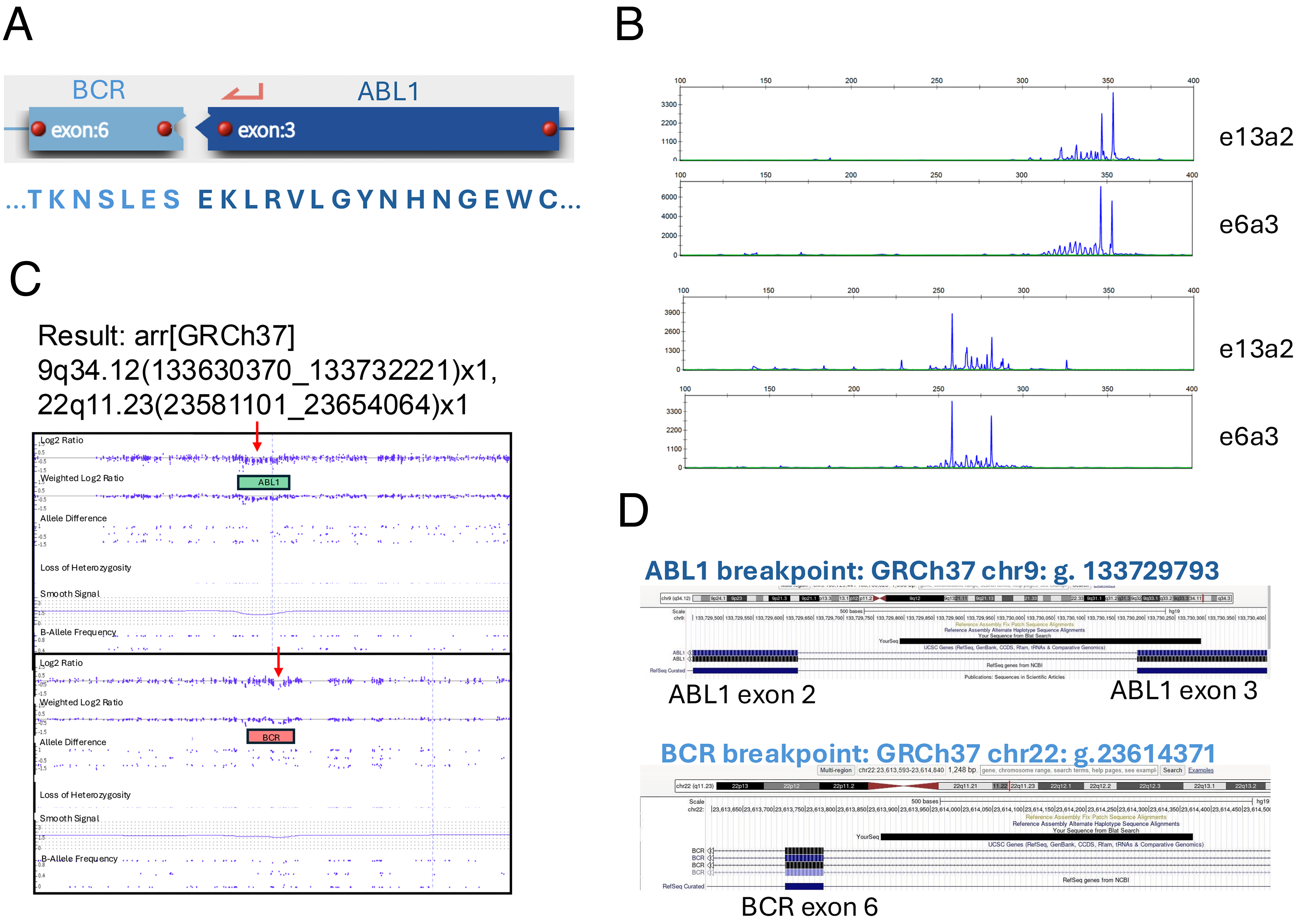


Supplemental Figure 1

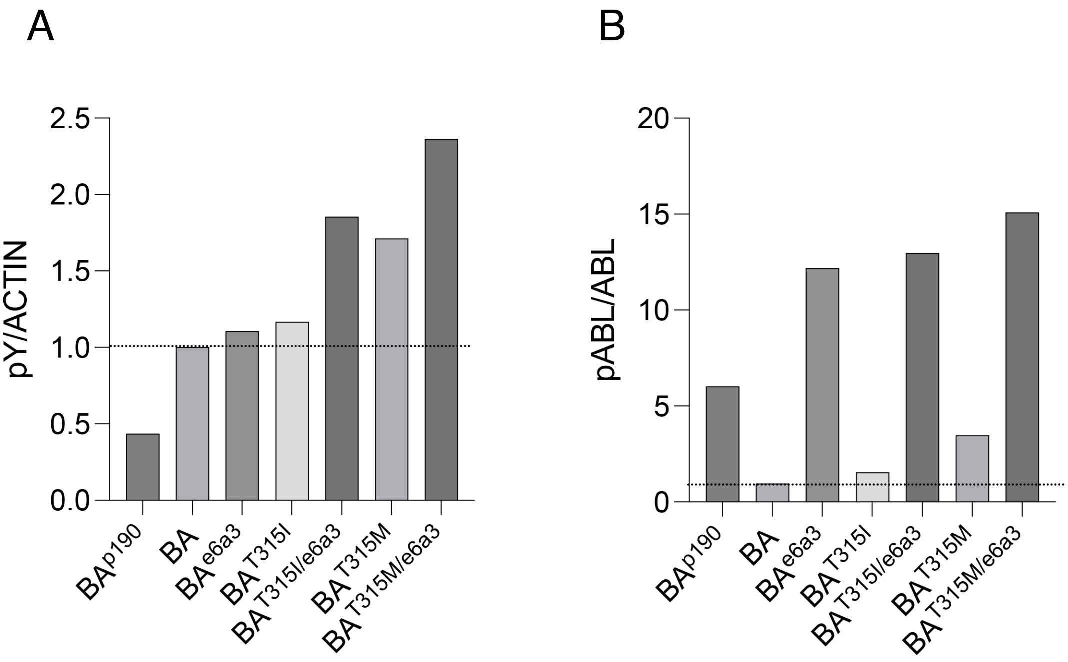


Supplemental Figure 2

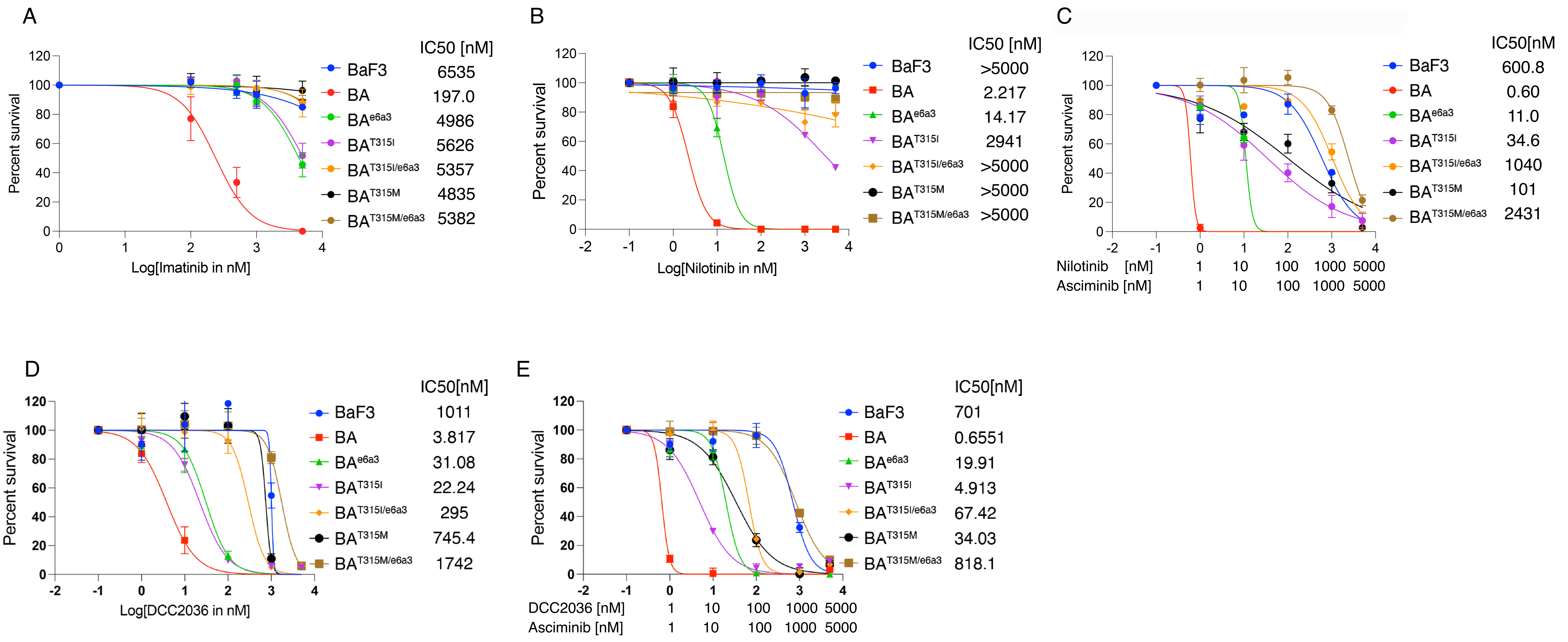


Supplemental Figure 3

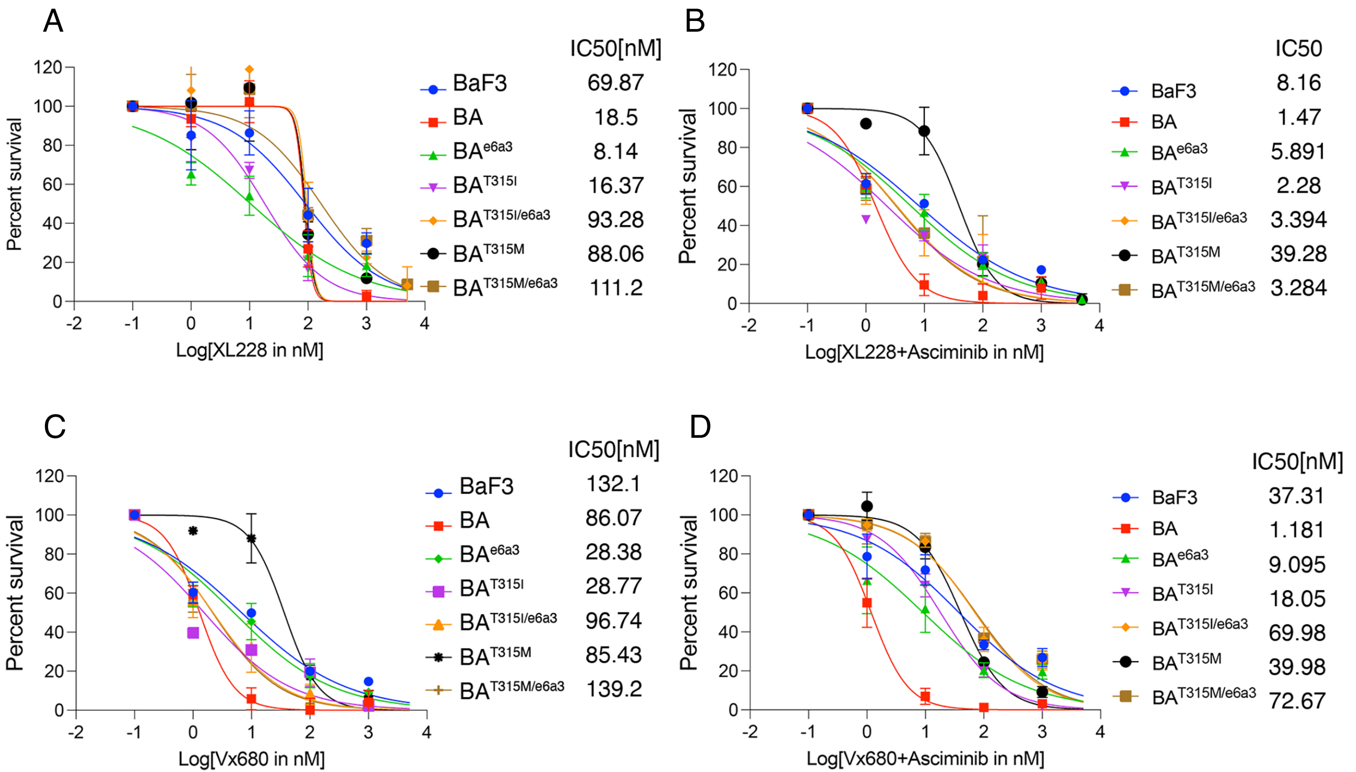


Supplemental Figure 4

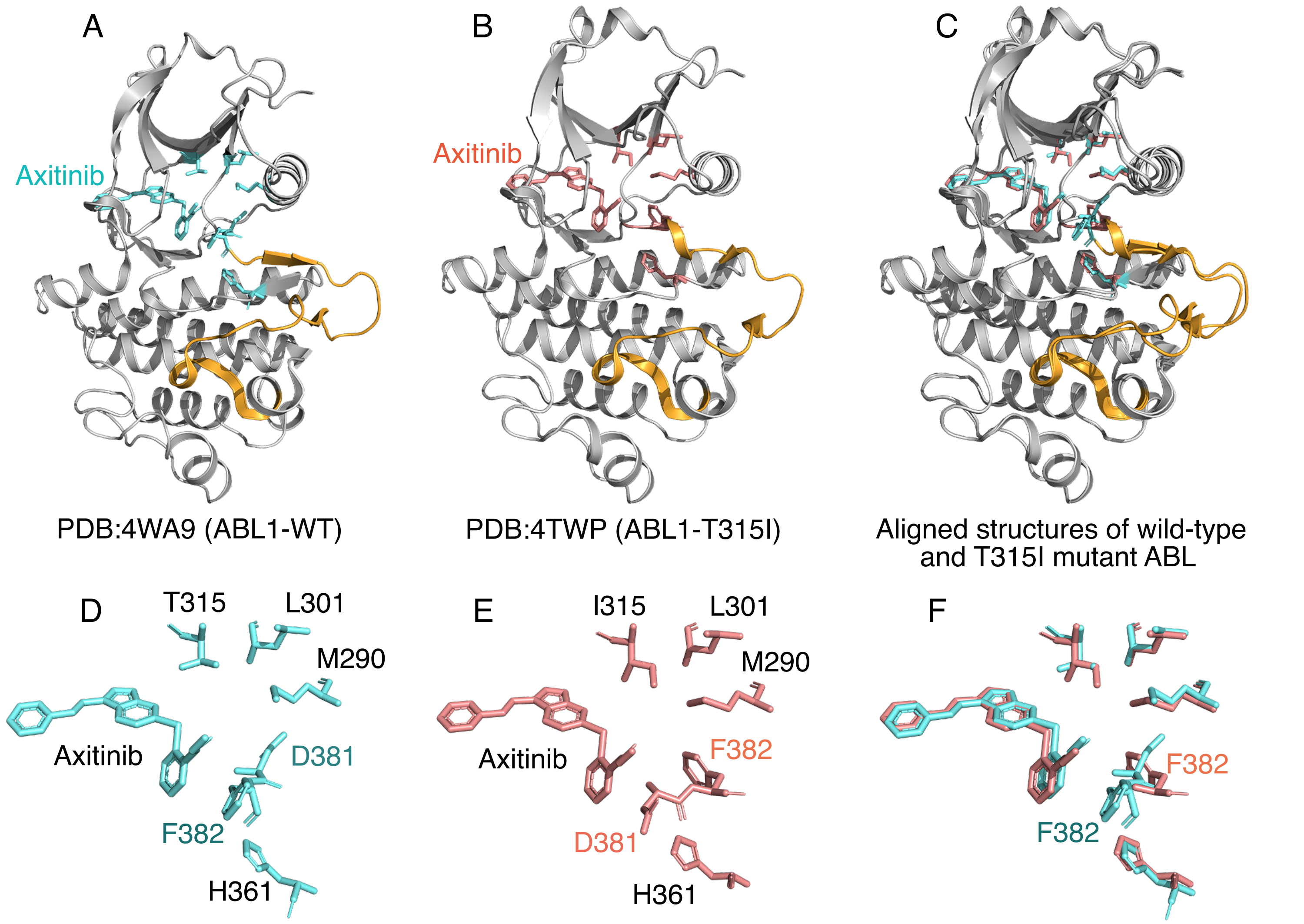


Supplemental Figure 5

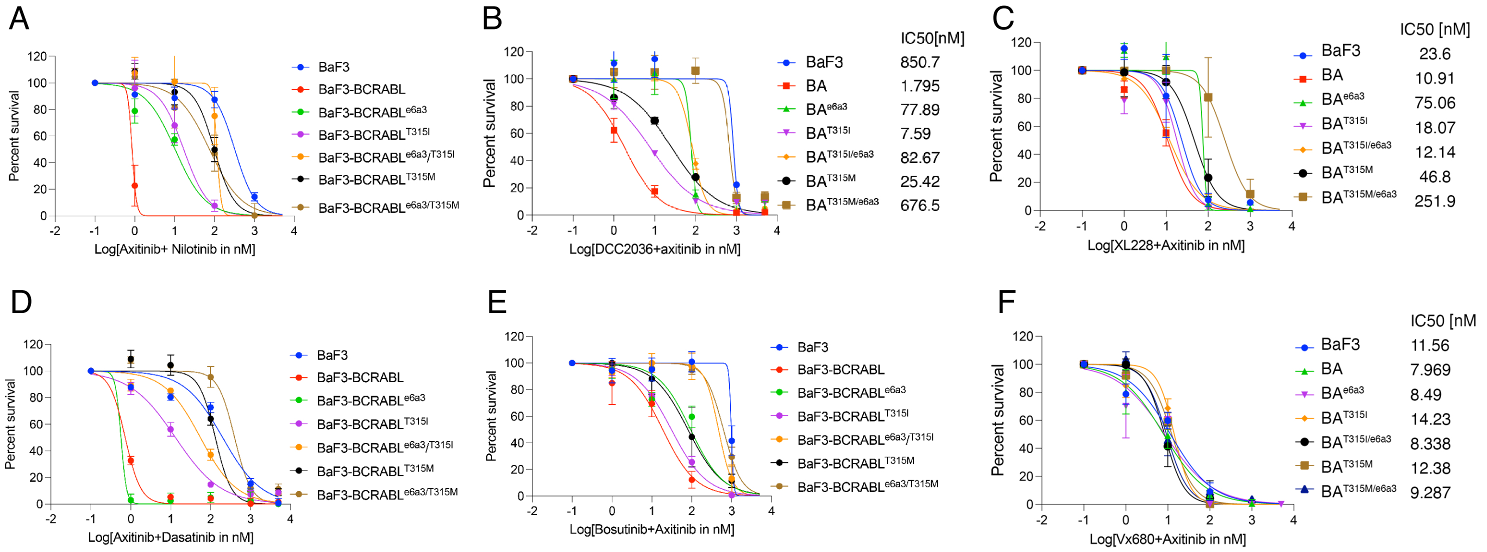


Supplemental Figure 6

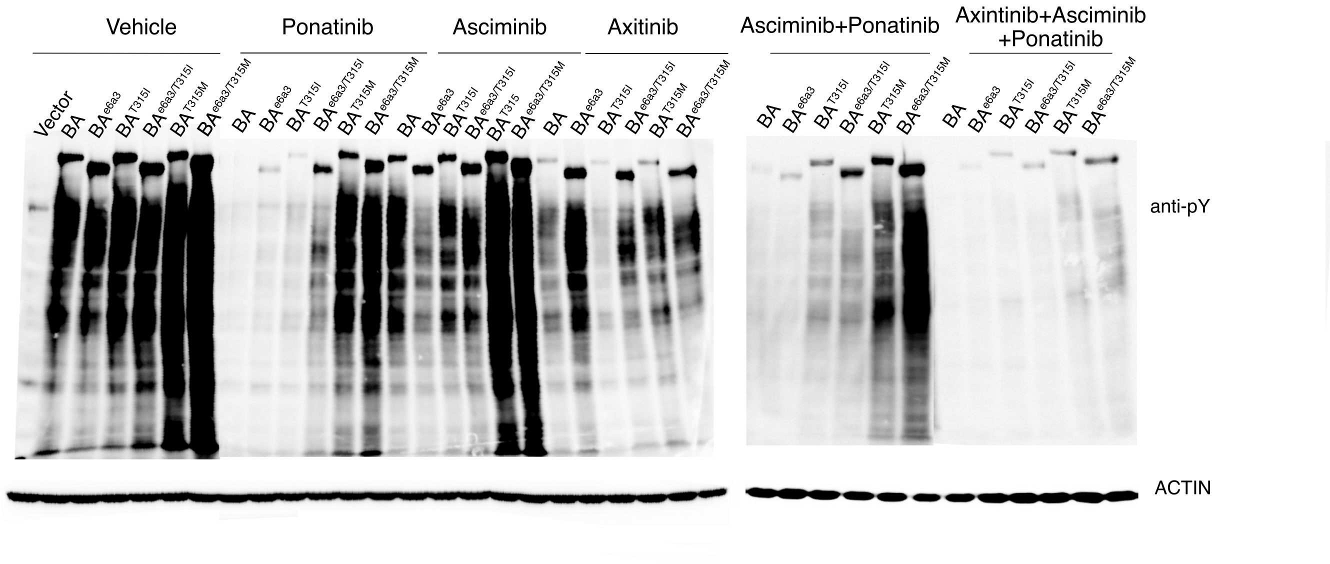


Supplemental Figure 7

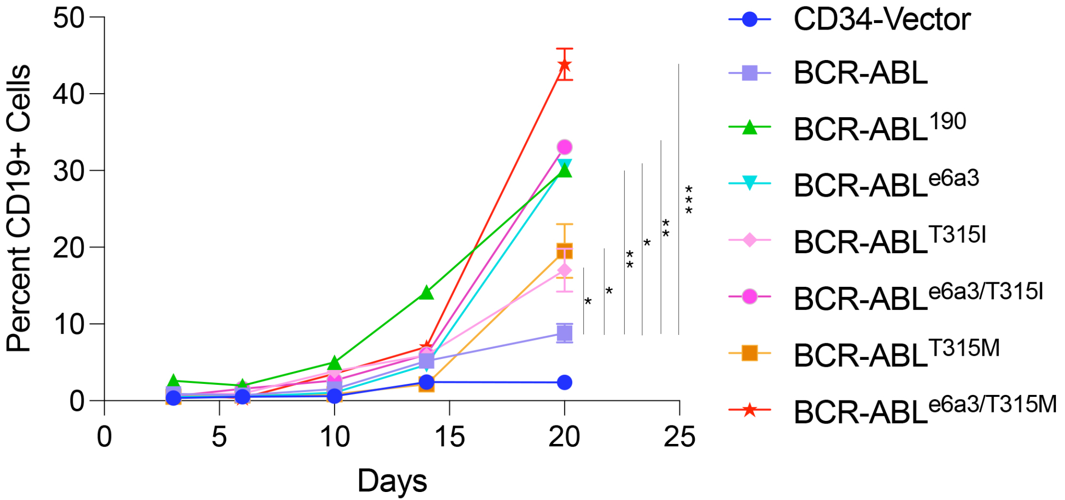


Supplemental Figure 8

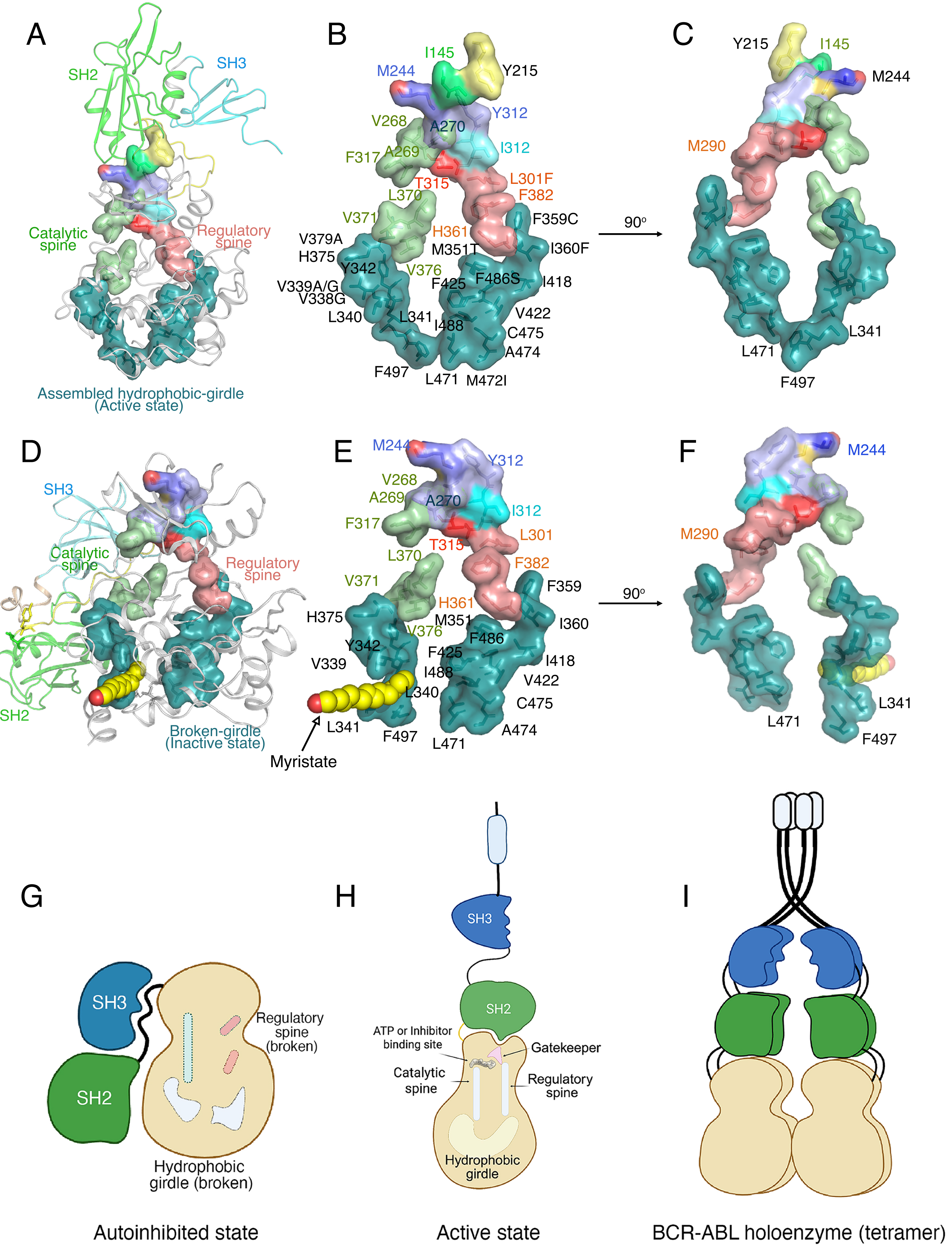


Supplemental Figure 9

### Supplemental Tables:

| Supplemental Table 1. Comparison of Genomic Findings in the Two Blast Crises | | |
|---|---|---|
| Feature / Gene | Initial e13a2 Blast Crisis | Blast Crisis e6a3 post asciminib + ponatinib |
| ***ABL1*** | c.749G>A (p.G250E)<br>VAF 24.1% | c.944C>T (p.T315I)<br>VAF 33.3% |
| ***BCORL1*** | c.3607+2T>C (splice variant)<br>VAF 4.3% | c.3607+2T>C (splice variant)<br>VAF 67.3% |
| ***DNMT3A*** | c.1627G>T (p.G543C)<br>VAF 52.6% | c.1627G>T (p.G543C)<br>VAF 45.7% |
| ***DNMT3A*** | Not detected | c.2014delG (p.V672Wfs*33)<br>VAF 2.3% |
| ***SF3B1*** | Not detected | c.1998G>T (p.K666N)<br>VAF 2.0% |
| ***IKZF1*** | No deletion detected | 1-copy deletion (7p) |
| ***RUNX1*** | No deletion detected | 1-copy deletion (21q) |

| Supplemental Table 2 | Asciminib | Ponatinib | Pon+Asc | Pon+Asc | Pon+Asc |
|---|---|---|---|---|---|
| | IC50[nM] | IC50[nM] | Das IC50 [nM] | Pon IC50 [nM] | CI |
| BaF3 | 4834 | 136.2 | 136.1 | 13.61 | 0.12808132 |
| BaF3-BCRABL | 0.6137 | 0.1734 | 0.1561 | 0.01561 | 0.34438188 |
| BaF3-BCRABL$^{e6a3}$ | 2549 | 1.761 | 0.2169 | 0.02169 | 0.01240196 |
| BaF3-BCRABL$^{T315I}$ | 30.07 | 1.108 | 0.548 | 0.0548 | 0.06768263 |
| BaF3-BCRABL$^{e6a3/T315I}$ | 4758 | 17.23 | 1.988 | 0.1988 | 0.01195584 |
| BaF3-BCRABL$^{T315M}$ | 394.1 | 81.64 | 2.641 | 0.2641 | 0.00993628 |
| BaF3-BCRABL$^{e6a3/T315M}$ | 8333 | 788.6 | 113.9 | 11.39 | 0.02811186 |

| Supplemental Table 3 | Imatinib | Nilotinib | Asciminib | Nilo+Asc | Nilo+Asc |
|---|---|---|---|---|---|
| | IC50[nM] | IC50[nM] | IC50[nM] | IC50[nM] | CI |
| BaF3 | 64757 | >5000 | 4834 | 600.8 | 0.24444631 |
| BaF3-BCRABL | 239.7 | 2.217 | 0.6137 | 0.6073 | 1.26350018 |
| BaF3-BCRABL$^{e6a3}$ | 4433 | 14.17 | 2549 | 11.06 | 0.78486119 |
| BaF3-BCRABL$^{T315I}$ | 5196 | 2941 | 30.07 | 34.65 | 1.16409298 |
| BaF3-BCRABL$^{e6a3/T315I}$ | 20347 | >5000 | 4758 | 1040 | 0.42657923 |
| BaF3-BCRABL$^{T315M}$ | 588614 | >5000 | 394.1 | 101 | 0.27648013 |
| BaF3-BCRABL$^{e6a3/T315M}$ | 24689 | >5000 | 8333 | 2431 | 0.77793167 |

| Supplemental Table 4 | DCC2036 | Asciminib | DCC2036+Asc | DCC2036+Asc |
|---|---|---|---|---|
| | IC50[nM] | IC50[nM] | IC50[nM] | CI |
| BaF3 | 1011 | 4834 | 701 | 0.83838738 |
| BaF3-BCRABL | 3.817 | 0.6137 | 0.6551 | 1.2390866 |
| BaF3-BCRABL$^{e6a3}$ | 31.08 | 2549 | 19.91 | 0.6484158 |

| | | | | |
|---|---|---|---|---|
| BaF3-BCRABL$^{T315I}$ | 22.24 | 30.07 | 4.913 | 0.38429371 |
| BaF3-BCRABL$^{e6a3/T315I}$ | 295 | 4758 | 67.42 | 0.24271219 |
| BaF3-BCRABL$^{T315M}$ | 745.4 | 394.1 | 34.03 | 0.13200198 |
| BaF3-BCRABL$^{e6a3/T315M}$ | 1742 | 8333 | 818.1 | 0.56780853 |

| Supplemental Table 5 | Dasatinib | Asciminib | Das+Asc | Das+Asc |
|---|---|---|---|---|
| | IC50[nM] | IC50[nM] | IC50[nM] | CI |
| BaF3 | 1015 | 4834 | 336.9 | 0.40161502 |
| BaF3-BCRABL | 1.155 | 0.6137 | 0.1571 | 0.39200558 |
| BaF3-BCRABL$^{e6a3}$ | 0.6701 | 2549 | 0.7058 | 1.05355252 |
| BaF3-BCRABL$^{T315I}$ | 1906 | 30.07 | 9.128 | 0.30834745 |
| BaF3-BCRABL$^{e6a3/T315I}$ | 1345 | 4758 | 1109 | 1.05761644 |
| BaF3-BCRABL$^{T315M}$ | 2083 | 394.1 | 6.446 | 0.01945083 |
| BaF3-BCRABL$^{e6a3/T315M}$ | 1130 | 8333 | 1184 | 1.18987329 |

| Supplemental Table 6 | Dasatinib | Ponatinib | Das+Pon | Das+Pon | Das+pon |
|---|---|---|---|---|---|
| | IC50[nM] | IC50[nM] | IC50 Das [nM] | IC50 Pon [nM] | CI |
| BaF3 | 1015 | 136.2 | 999.8 | 82.39 | 1.58994387 |
| BaF3-BCRABL | 1.155 | 0.1734 | 0.2251 | 0.02251 | 0.32470723 |
| BaF3-BCRABL$^{e6a3}$ | 0.6701 | 1.761 | 0.8979 | 0.08979 | 1.39093734 |
| BaF3-BCRABL$^{T315I}$ | 1906 | 1.108 | 12.44 | 1.243 | 1.12836791 |
| BaF3-BCRABL$^{e6a3/T315I}$ | 1345 | 17.23 | 97.61 | 9.734 | 0.63751735 |
| BaF3-BCRABL$^{T315M}$ | 2083 | 81.64 | 377 | 37.71 | 0.64289489 |
| BaF3-BCRABL$^{e6a3/T315M}$ | 1130 | 788.6 | 1529 | 180.4 | 1.58185717 |

| Supplemental Table 7 | Bosutinib | Asciminib | Bos+Asc | Bos+Asc |
|---|---|---|---|---|
| | IC50[nM] | IC50[nM] | IC50[nM] | CI |
| BaF3 | 4834 | 1105 | 522.1 | 0.58049448 |
| BaF3-BCRABL | 0.6137 | 46.79 | 1.214 | 2.00411094 |
| BaF3-BCRABL$^{e6a3}$ | 2549 | 21.11 | 100.7 | 4.80975675 |
| BaF3-BCRABL$^{T315I}$ | 30.07 | 1027 | 134.1 | 4.59016877 |
| BaF3-BCRABL$^{e6a3/T315I}$ | 4758 | 1235 | 999.3 | 1.01917502 |
| BaF3-BCRABL$^{T315M}$ | 394.1 | 1200 | 301.4 | 1.01594718 |
| BaF3-BCRABL$^{e6a3/T315M}$ | 8333 | 1128 | 1072 | 1.07899976 |

| Supplemental Table 8 | Bosutinib | Ponatinib | Bos+Pon | Bos+Pon | Bos+pon |
|---|---|---|---|---|---|
| | IC50[nM] | IC50[nM] | IC50 Das [nM] | IC50 Pon [nM] | CI |
| BaF3 | 136.2 | 1105 | 114.3 | 1098 | 1.83287221 |
| BaF3-BCRABL | 0.1734 | 46.79 | 0.3281 | 2.401 | 1.94347125 |
| BaF3-BCRABL$^{e6a3}$ | 1.761 | 21.11 | 1.446 | 13.2 | 1.44642043 |
| BaF3-BCRABL$^{T315I}$ | 1.108 | 1027 | 3.345 | 33.42 | 3.05149445 |

| | | | | | |
|---|---|---|---|---|---|
| BaF3-BCRABL$^{e6a3}$/$^{T315I}$ | 17.23 | 1235 | 70.3 | 339.5 | 4.35499165 |
| BaF3-BCRABL$^{T315M}$ | 81.64 | 1200 | 78.55 | 821.1 | 1.64640091 |
| BaF3-BCRABL$^{e6a3/T315M}$ | 788.6 | 1128 | 232.6 | 1000 | 1.1814779 |

| Supplemental Table 9 | VX680 | Asciminib | Vx+Asc | VX680+Asc |
|---|---|---|---|---|
| | | IC50[nM] | IC50 | CI |
| BaF3 | 132.1 | 4834 | 18.76 | 0.14589447 |
| BaF3-BCRABL | 86.07 | 0.6137 | 1.5 | 2.46161865 |
| BaF3-BCRABL$^{e6a3}$ | 28.38 | 2549 | 4.699 | 0.16741782 |
| BaF3-BCRABL$^{T315I}$ | 28.77 | 30.07 | 13.41 | 0.91206996 |
| BaF3-BCRABL$^{e6a3}$/$^{T315I}$ | 96.74 | 4758 | 47.23 | 0.49814228 |
| BaF3-BCRABL$^{T315M}$ | 85.43 | 394.1 | 27.87 | 0.3969501 |
| BaF3-BCRABL$^{e6a3/T315M}$ | 139.2 | 8333 | 47.61 | 0.34773929 |

| Supplemental Table 10 | XL228 | Asciminib | XL+Asc | XL228+Asc |
|---|---|---|---|---|
| | IC50 | IC50[nM] | IC50 | CI |
| BaF3 | 69.87 | 4834 | 6.76 | 0.09814954 |
| BaF3-BCRABL | 18.5 | 0.6137 | 1.299 | 2.1868856 |
| BaF3-BCRABL$^{e6a3}$ | 8.14 | 2549 | 5.002 | 0.61645865 |
| BaF3-BCRABL$^{T315I}$ | 16.37 | 30.07 | 1.549 | 0.14613745 |
| BaF3-BCRABL$^{e6a3}$/$^{T315I}$ | 93.28 | 4758 | 2.207 | 0.0241238 |
| BaF3-BCRABL$^{T315M}$ | 88.06 | 394.1 | 36.22 | 0.50321608 |
| BaF3-BCRABL$^{e6a3/T315M}$ | 111.2 | 8333 | 2.165 | 0.01972924 |

| Supplemental Table 11 | Nilotinib | Axitinib | Nilo+Ax | Nilo+Ax |
|---|---|---|---|---|
| | IC50[nM] | IC50[nM] | IC50[nM] | CI |
| BaF3 | 5000 | 911 | 227.4 | 0.2950958 |
| BaF3-BCRABL | 2.217 | 141 | 0.8297 | 0.3801289 |
| BaF3-BCRABL$^{e6a3}$ | 14.17 | 778.1 | 9.856 | 0.7082207 |
| BaF3-BCRABL$^{T315I}$ | 2941 | 12.5 | 5.298 | 0.4256414 |
| BaF3-BCRABL$^{e6a3}$/$^{T315I}$ | 5000 | 410 | 150.1 | 0.3961176 |
| BaF3-BCRABL$^{T315M}$ | 5000 | 215 | 25.69 | 0.1246264 |
| BaF3-BCRABL$^{e6a3/T315M}$ | 5000 | 486 | 366.7 | 0.8278667 |

| Supplemental Table 12 | DCC2036 | Axitinib | DCC2036+Axi | DCC2036+Ax |
|---|---|---|---|---|
| | IC50[nM] | IC50[nM] | IC50[nM] | CI |
| BaF3 | 1011 | 911 | 850.7 | 1.7752531 |
| BaF3-BCRABL | 3.817 | 141 | 1.795 | 0.4829951 |
| BaF3-BCRABL$^{e6a3}$ | 31.08 | 778.1 | 77.89 | 2.6062161 |
| BaF3-BCRABL$^{T315I}$ | 22.24 | 12.5 | 7.59 | 0.948477 |

| | | | | |
|---|---|---|---|---|
| BaF3-BCRABL$^{e6a3/T315I}$ | 295 | 410 | 82.67 | 0.4818714 |
| BaF3-BCRABL$^{T315M}$ | 745.4 | 215 | 25.42 | 0.1523351 |
| BaF3-BCRABL$^{e6a3/T315M}$ | 1742 | 486 | 676.5 | 1.780322 |

| Supplemental Table 13 | Dasatinib | Axitinib | Das+Ax | Das+Ax |
|---|---|---|---|---|
| | IC50[nM] | IC50[nM] | IC50[nM] | CI |
| BaF3 | 1015 | 911 | 200.3 | 0.4172082 |
| BaF3-BCRABL | 1.155 | 141 | 0.7242 | 0.6321492 |
| BaF3-BCRABL$^{e6a3}$ | 0.6701 | 778.1 | 0.5669 | 0.8467218 |
| BaF3-BCRABL$^{T315I}$ | 1906 | 12.5 | 13.14 | 1.058094 |
| BaF3-BCRABL$^{e6a3/T315I}$ | 1345 | 410 | 55.45 | 0.1764707 |
| BaF3-BCRABL$^{T315M}$ | 2083 | 215 | 131.1 | 0.6727055 |
| BaF3-BCRABL$^{e6a3/T315M}$ | 1130 | 486 | 399.5 | 1.1755563 |

| Supplemental Table 14 | Bosutinib | Axitinib | Bos+Ax | Bos+Ax |
|---|---|---|---|---|
| | IC50[nM] | IC50[nM] | IC50[nM] | CI |
| BaF3 | 1105 | 911 | 977.8 | 1.9582129 |
| BaF3-BCRABL | 46.79 | 141 | 18.73 | 0.5331361 |
| BaF3-BCRABL$^{e6a3}$ | 21.11 | 778 | 93.33 | 4.5410889 |
| BaF3-BCRABL$^{T315I}$ | 1027 | 12.5 | 30.83 | 2.4964195 |
| BaF3-BCRABL$^{e6a3/T315I}$ | 1235 | 410 | 468.7 | 1.5226849 |
| BaF3-BCRABL$^{T315M}$ | 1200 | 215 | 80.21 | 0.4399114 |
| BaF3-BCRABL$^{e6a3/T315M}$ | 1128 | 486 | 622.4 | 1.8324315 |

| Supplemental Table 15 | VX680 | Axitinib | VX+Axi | VX680+Axi |
|---|---|---|---|---|
| | | IC50[nM] | IC50 | CI |
| BaF3 | 132.1 | 911 | 11.56 | 0.1001988 |
| BaF3-BCRABL | 86.07 | 141 | 7.969 | 0.1491052 |
| BaF3-BCRABL$^{e6a3}$ | 28.38 | 778.1 | 8.49 | 0.3100655 |
| BaF3-BCRABL$^{T315I}$ | 28.77 | 12.5 | 14.23 | 1.6330124 |
| BaF3-BCRABL$^{e6a3/T315I}$ | 96.74 | 410 | 8.338 | 0.1065264 |
| BaF3-BCRABL$^{T315M}$ | 85.43 | 215 | 12.38 | 0.2024954 |
| BaF3-BCRABL$^{e6a3/T315M}$ | 139.2 | 486 | 9.287 | 0.085826 |

| Supplemental Table 16 | XL228 | Axitinib | XL+Axi | XL228+Axi |
|---|---|---|---|---|
| | IC50 | IC50[nM] | IC50 | CI |
| BaF3 | 69.87 | 911 | 23.6 | 0.3636757 |
| BaF3-BCRABL | 18.5 | 141 | 10.91 | 0.6671056 |
| BaF3-BCRABL$^{e6a3}$ | 8.14 | 778.1 | 75.06 | 9.317596 |
| BaF3-BCRABL$^{T315I}$ | 16.37 | 12.5 | 18.07 | 2.5494485 |
| BaF3-BCRABL$^{e6a3/T315I}$ | 93.28 | 410 | 12.14 | 0.1597556 |

| | | | | |
|---|---|---|---|---|
| BaF3-BCRABL$^{T315M}$ | 88.06 | 215 | 46.8 | 0.7491302 |
| BaF3-BCRABL$^{e6a3/T315M}$ | 111.2 | 486 | 251.9 | 2.7836005 |

| Supplemental Table 17 | Asciminib | Ponatinib | Axitinib | Asc+Pon | Axi+Pon | Pon+Ax+As |
|---|---|---|---|---|---|---|
| | IC50[nM] | IC50[nM] | IC50[nM] | IC50[nM] | IC50[nM] | IC50[nM] |
| BaF3 | 4834 | 136.2 | 911 | 136.1 | 897.1 | 1344 |
| BaF3-BCRABL | 0.6137 | 0.1734 | 141 | 0.1561 | 1.007 | 0.5618 |
| BaF3-BCRABL$^{e6a3}$ | 2549 | 1.761 | 778.1 | 0.2169 | 63.67 | 0.571 |
| BaF3-BCRABL$^{T315I}$ | 30.07 | 1.108 | 12.5 | 0.548 | 10.76 | 0.5793 |
| BaF3-BCRABL$^{e6a3/T315I}$ | 4758 | 17.23 | 410 | 1.988 | 764 | 0.5925 |
| BaF3-BCRABL$^{T315M}$ | 394.1 | 81.64 | 215 | 2.641 | 117.7 | 7.87 |
| BaF3-BCRABL$^{e6a3/T315M}$ | 8333 | 788.6 | 486 | 113.9 | 921.5 | 213.6 |
